# Polymorphisms affecting expression of the vaccine antigen factor H binding protein influence invasiveness of Neisseria meningitidis

**DOI:** 10.1101/2021.01.08.21249443

**Authors:** Sarah G. Earle, Mariya Lobanovska, Hayley Lavender, Changyan Tang, Rachel M. Exley, Elisa Ramos-Sevillano, Douglas Browning, Vasiliki Kostiou, Odile B. Harrison, Holly B. Bratcher, Gabriele Varani, Christoph M. Tang, Daniel J. Wilson, Martin C. J. Maiden

## Abstract

Many bacterial diseases are caused by organisms that ordinarily are harmless components of the human microbiome. Effective interventions against these conditions requires an understanding of the processes whereby symbiosis or commensalism breaks down. Here, we performed bacterial genome-wide association studies (GWAS) of *Neisseria meningitidis*, a common commensal of the human respiratory tract despite being a leading cause of meningitis and sepsis. GWAS discovered single nucleotide polymorphisms (SNPs) and other bacterial genetic variants associated with invasive meningococcal disease (IMD) *versus* carriage in several loci across the genome, revealing the polygenic nature of this phenotype. Of note, we detected a significant peak around *fHbp*, which encodes factor H binding protein (fHbp); fHbp promotes bacterial immune evasion of human complement by recruiting complement factor H (CFH) to the meningococcal surface. We confirmed the association around fHbp with IMD in a validation GWAS, and found that SNPs identified in the validation affecting the 5’ region of *fHbp* mRNA alter secondary RNA structures, increase fHbp expression, and enhance bacterial escape from complement-mediated killing. This finding mirrors the known link between complement deficiencies and CFH variation with human susceptibility to IMD, highlighting the central importance of human and bacterial genetic variation across the fHbp:CFH interface in IMD susceptibility, virulence, and the transition from carriage to disease.

---

Many members of the human microbiota are capable of causing disease in certain circumstances. Given the complexity of relationships between commensal and symbiotic bacteria and their hosts, there are many reasons why interactions become disrupted, including genetic polymorphisms and phenotypic changes in hosts and infecting microbes. These diseases present both an evolutionary puzzle, as disease is often a dead-end for transmission, and an epidemiological challenge, as the aetiological agent circulates widely undetected, striking seemingly at random. An important example of such an ‘accidental’ pathogen is *Neisseria meningitidis*, a common coloniser of the human oropharynx [1, 2], which occasionally causes devastating invasive meningococcal disease (IMD).

The molecular mechanisms involved in the transition of asymptomatic colonisation to IMD remain poorly defined despite extensive research. Invasive meningococci almost invariably express specific capsular polysaccharides (serogroups A, B, C, X or W) and belong to certain lineages (the ‘hypervirulent’ meningococci). On the other hand, complement deficiencies are a well-known human risk factor [3], but do not explain the overwhelming majority of cases; however, host susceptibility to IMD is linked to genetic variation in the locus encoding the negative regulator of the complement system, complement factor H (CFH) [4, 5]. *N. meningitidis* expresses factor H binding protein (fHbp), an important vaccine antigen, which binds CFH with high affinity, and promotes evasion of complement-mediated killing [6, 7].

We explored the relationship between meningococcal genetic variation and IMD in two genome wide association studies (GWAS) encompassing 1,556 genomes. Initially, we compared *N. meningitidis* isolates from an outbreak of 52 cases of IMD and 209 carriers in the Czech Republic [8, 9, 10], in which a preponderance of disease isolates belonged to the hypervirulent clonal complex ST-11 (cc; O.R. 3.4, 95% CI 1.7-7.1, Wald test *p*=10^−3.90^) [11, 12] (**Figure 1A, Supplementary Figure 1**). Heritability was substantial, with 36.5% (95% CI 15.9-57.0%) of the variability in case-control status attributable to bacterial genetics. GWAS identified 17 loci harbouring variants associated with carriage *versus* disease, after controlling for strain-to-strain differences using a linear mixed model [13]. In total, we tested for associations at 156,804 SNPs mapped to an ST-11 complex reference genome [14] and 7,806,583 31-nucleotide sequence fragments (kmers) that capture variation missed by reference-based SNP calling [15]. After Bonferroni correction for unique phylopatterns [16], we found significant associations in seven SNPs (*p*<10^−6.20^) and 465 kmers (*p*<10^−6.79^) across the genome (**Figure 1B and C**).

**Figure 1.**
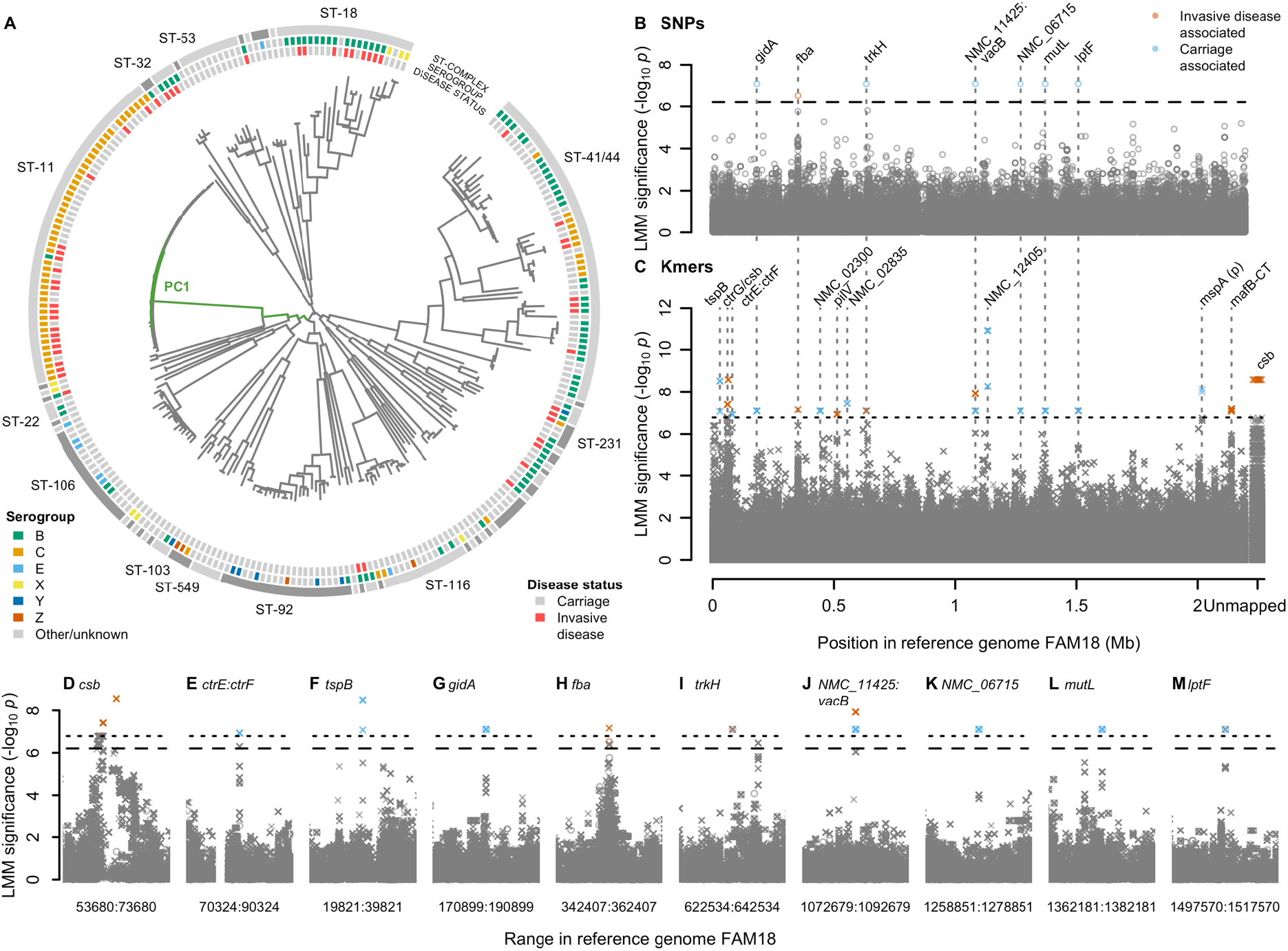
**(A)** Phylogeny of 261 *N. meningitidis* strains sampled from the Czech Republic in 1993 shows a strong strain association between invasive disease and the ST-11 complex. Clonal complexes are shown in the outer grey ring. Serogroups are shown on the next ring inwards. Disease status is shown on the next ring, invasive disease (red, n = 209) or carriage (grey, n = 52). Branches of the phylogeny most correlated with the significantly associated PC 1 are coloured in green. **(B-J)** SNPs and kmers associated with carriage *vs*. invasive disease in the 261 isolates. Significant SNPs and kmers are coloured by the LMM estimated direction of effect. Bonferroni-corrected significance thresholds are shown by black dashed (SNPs) and dotted (kmers) lines. Gene names separated by colons indicate intergenic regions. FAM18 reference genome gene name prefixes have been shortened from NMC_RS to NMC_. **(B)** Each point represents a SNP aligned to the reference genome FAM18. **(C)** Each point represents the left-most position of a kmer in the reference genome FAM18 based on mapping and BLAST alignments. Unmapped kmers are plotted to the right of the Manhattan plot. **(D-M)** Close ups of genes containing significant SNPs (circles) or kmers (crosses) +/-10kb, SNPs and kmers are shown.

GWAS identified variation in several genes associated with known virulence factors including capsule and the meningococcal disease-associated (MDA) island (**Figure 1D-F, Supplementary Text 1**) [17, 18, 19, 20]. In summary, 21 kmers in the gene *csb* encoding the capsule polysialyltransferase were associated with elevated disease risk (*p*=10^−8.58^) and mapped to a poly-A tract in the coding region which mediates ON:OFF switching of capsule expression [21], capturing the capsule ON status (**Supplementary Figure 2**). Nine kmers which tagged the consensus haplotype across three SNPs upstream of *ctrF* (involved in capsule export), between the predicted -10 and -35 sequences of the *ctrF* promotor, were more frequent in carriage (*P*=10^−6.93^) and may affect transcription (**Supplementary Figure 3**). Within the MDA island, nine carriage-associated kmers tagged SNPs in the IgG binding domain of *tspB* (*p*=10^−8.51^; **Supplementary Figure 4**) [19, 20]. Several other loci contained significant hits (**Supplementary Table 1**), including a band of SNPs in perfect linkage disequilibrium in six genes (*p*=10^−7.10^; **Figure 1 G-M**).

**Figure 2.**
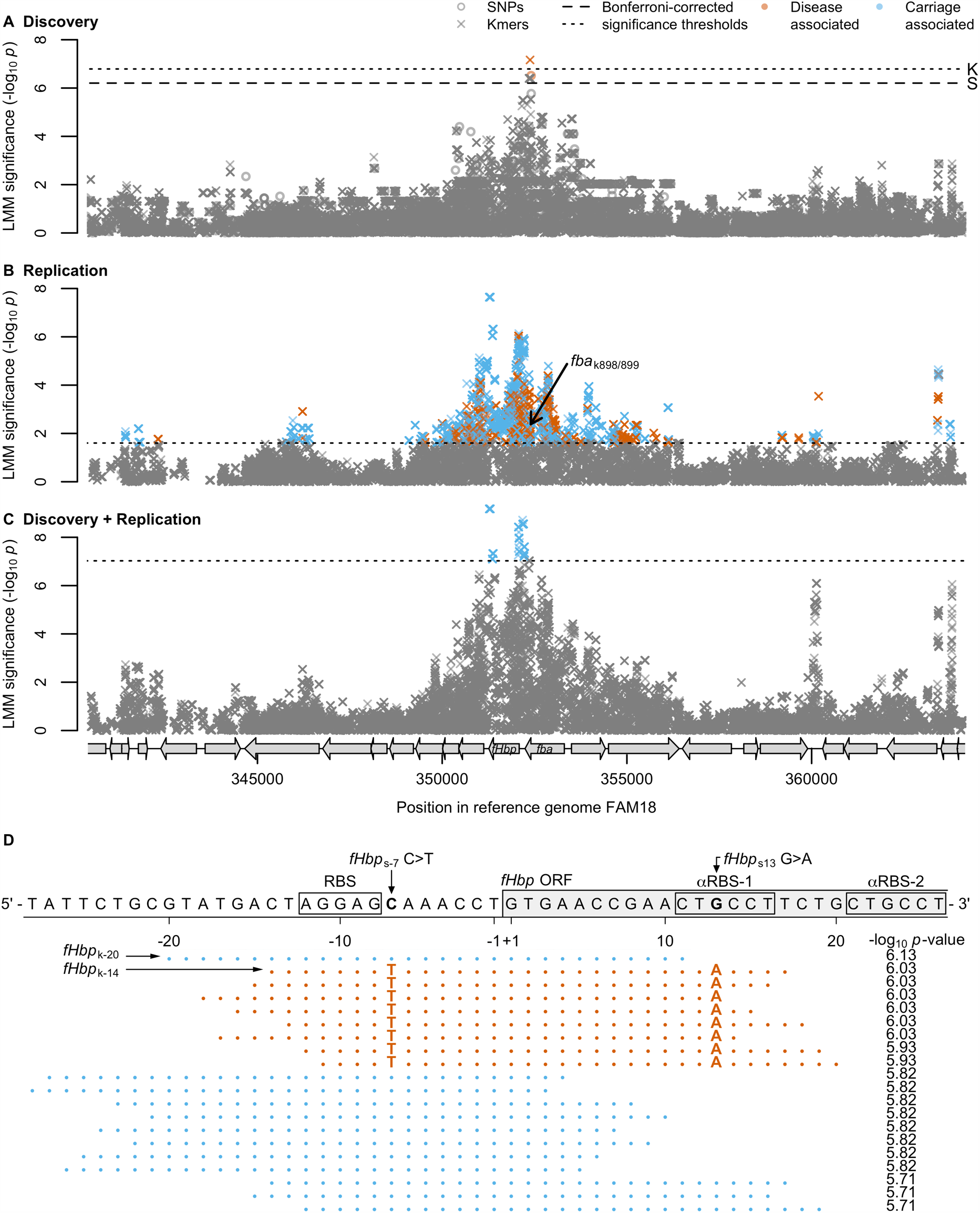
**(A-C)** SNPs and kmers in the *fba*-*fHbp* operon plus the surrounding 10kb. Open circles represent SNPs and crosses represent left-most kmer mapping positions. Significant SNPs and kmers are coloured by the LMM estimated direction of effect. Bonferroni-corrected significance thresholds are shown by black dashed lines (SNPs) or black dotted lines (kmers). Grey arrows represent coding sequences in the reference genome FAM18. **(A)** The discovery 261 isolates sampled from the Czech Republic in 1993. **(B)** Kmers above 1% minor allele frequency (MAF) in 1,295 ST-41/44 complex replication study genomes. The black arrow points to the position and significance of the two kmers in the gene *fba* that were significant in the discovery sample collection. **(C)** Kmers above 1% (MAF) in the discovery 261 Czech Republic sample collection plus the 1,295 ST-41/44 complex replication study genomes. **(D)** The 20 most significant kmers in the replication study surrounding the *fHbp* start codon. The FAM18 reference genome is shown for positions 352112-352059. The start of the open reading frame (ORF), the ribosomal binding site (RBS) and two putative anti-RBS sites are annotated. Kmer sequences are depicted by dots where they are the same as the reference, and by their base where they differ. Blue kmers are estimated to be associated with carriage, and dark orange kmers with invasive disease. The kmer -log_10_ *p*-values are annotated. Of the top 20 kmers in this region, the carriage-associated kmers were identical to FAM18, and the disease-associated kmers contained two annotated SNPs, a T at fHbp position -7, 1 bp away from the RBS, and an A at position 13 within the putative anti-RBS-1 sequence.

**Figure 3.**
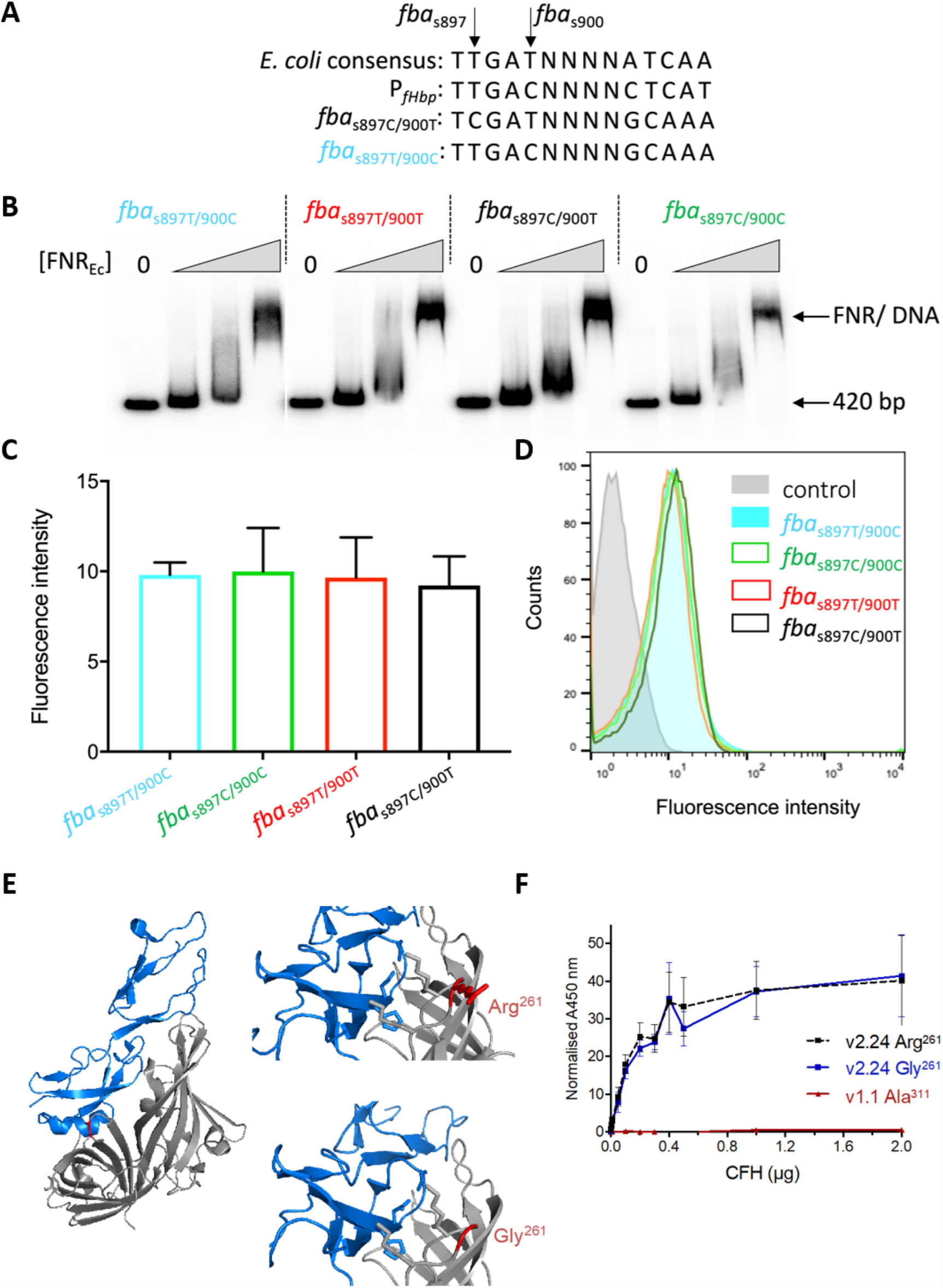
SNPs in *fba* do not alter FNR binding or fHbp surface expression. **(A)** Sequences of SNPs in *fba* aligned with the consensus *E. coli* FNR binding sequence and the known FNR site upstream of *fHbp*. **(B)** EMSA of 420 bp upstream of *fHbp* (including the known FNR binding site) with increasing concentrations of FNR (0, 0.75, 1.5, 3 µM). **(C)** fHbp was detected on the surface of bacteria with different SNPs (indicated) by flow cytometry using α-fHbp pAbs. Geometric mean fluorescence was used to compare fHbp levels across the samples. Error bars show SEM, significance analysed by two-way ANOVA showed no statistical difference between the strains. **(D)** Representative flow cytometry histograms; strains indicated; control (grey filled area), no primary pAb. **(E)** Side chains of Arg^261^ and Gly^261^ (red) of fHbp (grey) shown with CCPs 6 and 7 of CFH (blue) and threaded onto fHbp (v3.28, PDB:4AYI); figures generated in PyMOL. **(F)** Binding of fHbps to CFH by ELISA; a non-functional fHbp (v1.1 Ala^311^) was included as a control; error bars, SD, n = 3.

**Figure 4.**
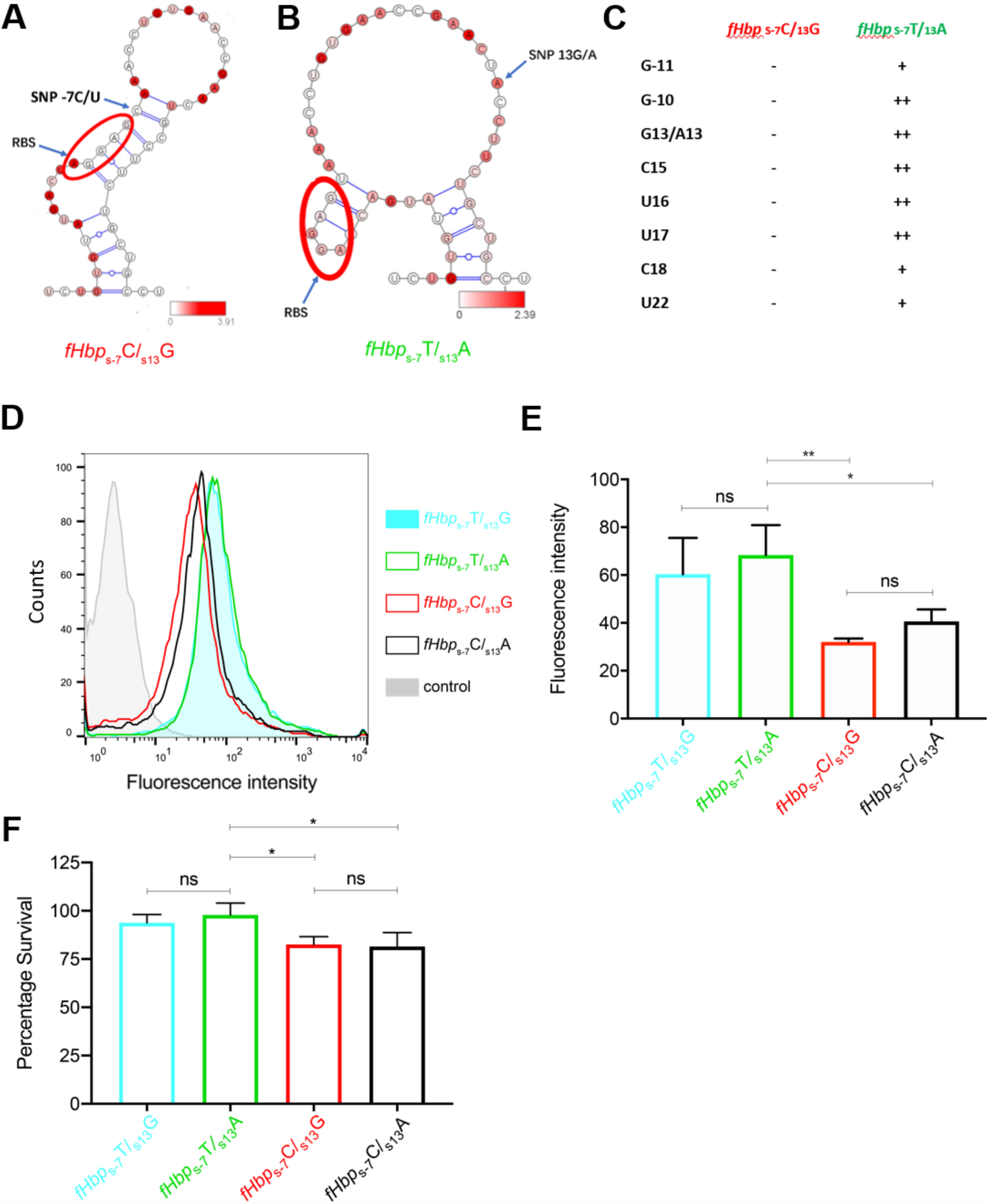
Secondary structure of the RNA structure at 37^°^C around the RBS calculated using RNAstructure and SHAPE reactivity data of **(A)** *fHbp*_s-7_C/*fHbp*_s13_G and **(B)** *fHbp*_s-7_T/*fHbp*_s13_A; SHAPE reactivity data are mapped on the RNA structure and colour coded by intensity as shown on the bars; the RBS is circled in red **(C)** Nucleotides with reactivity are listed in the table as strong (++), medium (+) and weak (-). Representative flow cytometry histograms and geometric mean fluorescence **(D, E)** of surface fHbp on *N. meningitidis* with SNPs *fHbp*_s-7_T/*fHbp*_s13_G (blue filled area), *fHbp*_s-7_T/*fHbp*_s13_A (solid green line), *fHbp*_s-7_C/*fHbp*_s13_G (solid red line) or *fHbp*_s-7_C/*fHbp*_s13_A (solid black line). Bacteria were grown at temperatures indicated, and fHbp detected with anti-fHbp pAb; control (grey filled), bacteria with no primary antibody. **(F)** Serum sensitivity assays of *N. meningitidis* strains with SNPs as indicated. Error bars show SD (n=3), * *p*<0.05, ** *p*<0.01 (n=3, two-way ANOVA).

Notably, our analysis identified novel signals in a region of elevated significance within the *fba-fHbp* operon (**Figure 2A**). *fba* encodes fructose-1, 6-bisphosphate aldolase (Fba) which functions in carbon metabolism and cell adhesion [22, 23] while *fHbp* encodes fHbp. The *fba-fHbp* signals were i) independent of the six other genome-wide significant SNPs, ii) physically localised in a single region (unlike other polymorphisms), and iii) displayed a significant decay of signal around a prominent peak, characteristic of an authentic association (**Figure 1D-M, Supplementary Figure 5**). The significant IMD-associated SNP (*P*=10^−6.51^) occurred at high frequency in the sample (58.7% invasive cases, 17.2% carriers), and explained 10% of sample heritability. Therefore, while several signals were detected across the genome, the association at *fba-fHbp* was of particular interest.

The significant SNP in the *fba-fHbp* locus occurred at nucleotide 900 of *fba* (*fba*_S900_, *P*=10^−6.51^), near the 3’ end of *fba*, as did two kmers spanning this SNP commencing at nucleotides 898 and 899 of *fba* (*fba*_K898_ and *fba*_K899_; *p*=10^−7.16^). There was a genome-wide significant enrichment in the rest of the *fba-fHbp* operon as a whole (adjusted harmonic mean *p*=10^−1.72^). The two kmers spanned protein-coding nucleotides 898-929, tagging the *fba*_S900_ SNP and two others: *fba*_S912_ (*p*=10^−2.25^) and *fba*_S913_ (*p*=10^−2.25^) (**Figure 2A, Supplementary Figure 6**). The peak signal of association therefore coincided with *fba*_S900_, which was in tight linkage disequilibrium with a neighbouring SNP *fba*_S897_ (*P*=10^−5.77^, *r*^2^ = 0.98). *fba*_S900_ and *fba*_S897_ both cause synonymous substitutions, located 323 and 326 bp upstream of the *fHbp* start codon, respectively.

A replication study was undertaken to test the association of *fba*_S900_ with IMD using genomes of an extended set of 1,295 clonal complex ST-41/44 meningococci, comprising 1046 IMD and 249 carriage isolates (available at https://PubMLST.org/neisseria [24]) (**Supplementary Figure 7**). Clonal complex ST-41/44 strains are the leading cause of IMD world-wide [25], and are polymorphic at *fba*_S900_. Analysing a single clonal complex mitigates confounding due to heterogeneous sampling across diverse lineages [26, 27]. After Bonferroni correction for two candidate kmers (*p*<10^−1.60^), the IMD-associated signal from kmers *fba*_K898_ and *fba*_K899_ was replicated in the ST-41/44 complex isolates (*p*=10^−2.37^), with the direction of the effect replicated (β=0.16). Moreover, the general enrichment in significance in *fba*-*fHbp* was replicated (adjusted harmonic mean *p*=10^−1.96^).

We explored possible effects of the synonymous SNPs in *fba* on the expression of *fHbp*, which can be translated from a bicistronic *fba-fHbp* mRNA or from a *fHbp*-specific promoter [28]. Expression of *fHbp* can be regulated by FNR binding to sequences 80 bp upstream of the start codon [28]. We noticed that the synonymous IMD-associated substitutions at *fba*_S897_ and *fba*_S900_ form a motif resembling an FNR box 314 bp upstream of *fHbp* (**Figure 3A, Supplementary Figure 6 and Text 2**). Electrophoresis mobility shift assays (EMSA) demonstrated binding of a constitutively active version of FNR to the known FNR site but not to sequences within *fba*, irrespective of the SNP sequence (**Figure 3B, Supplementary Figure 8**). Furthermore, there was no detectable difference in fHbp expression by four isogenic strains covering all combinations of the SNPs *fba*_S897_C/T and *fba*_S900_T/C in an ST-41/44 complex strain (**Figure 3C-D, Supplementary Text 2**).

Therefore, we considered whether other variants in the *fba-fHbp* region could be driving the signals of association as a total of 1,346 kmers in *fba-fHbp* were more significant in the replication study than the candidate kmers, *fba*_K898_ and *fba*_K899_ (**Figure 2B-D**). The most significant kmers above 1% minor allele frequency in the replication study were those starting at *fHbp* nt -20 and -14 (henceforth *fHbp*_K-20_ and *fHbp*_K-14_, respectively *p*<10^−5.93^), at nt 686 (*fHbp*_K686_, *p*<10^−6.04^), and nt 752 (*fHbp*_K752_, *p*<10^−7.64^) relative to the first base of the start codon. The SNPs tagged by these kmers were: i) *fHbp*_S-7_C/T in the 5’-untranslated region (5’-UTR) of *fHbp* adjacent to the ribosome binding site (RBS, *p*=10^−6.13^); ii) *fHbp*_S13_G/A encoding an Ala^5^Thr substitution in *fHbp* near a previously identified anti-RBS (α-RBS) site (*p*=10^−6.03^) [29]; iii) *fHbp*_S781_A/G which leads to an Arg^261^Gly substitution in fHbp adjacent to the CFH binding site (*p*=10^−7.64^); and iv) *fHbp*_S700_G/A causing a Gly^234^Ser substitution distant from the site of CFH (*p*=10^−6.32^) (**Figure 3E, Supplementary Figures 9-11**).

The IMD-associated SNP *fHbp*_S781_G removes a charged side chain (on Arg^261^) which could affect interactions with CFH (**Figure 3E**). We generated proteins with Arg^261^ or Gly^261^ in v2.24 fHbp, the allele most significantly associated with IMD (*p*=10^−3.23^, **Supplementary Figure 12**), and assessed fHbp:CFH binding by ELISA. However, we found no evidence that *fHbp*_S781_ altered binding to CFH (**Figure 3F**).

To explore an alternative mechanism, we examined the effect of the SNPs around the RBS and α-RBS sequence using SHAPE chemistry to probe the RNA secondary structure using a 183 nt RNA encompassing the RBS at position -8 to -12 (relative to the translation start site), and *fHbp*_S-7_T/C and *fHbp*_S13_A/G. The secondary structure model based on SHAPE reactivity data of *fHbp*_S-7_C/_S13_G at 37^°^C (**Figure 4A**) is consistent with the RBS being base-paired and masked through the formation of a relatively long imperfect helix of 11 base pairs that includes both anti-RBS sequences 1 (αRBS-1) and 2 (αRBS-1) [29]; the polymorphic sites in the carriage associated *fHbp*_S-7_C/_S13_G structure form a G:C base pair at the top of the helix. However, the local RNA structure of the IMD-associated *fHbp*_S-7_T/_S13_A shows significant differences (**Figure 4B-C**), with the 6 bp structure around the RBS being much more open and accessible (**Supplementary Figures 13-14** for SHAPE analysis and predicted RNA structures at 30^°^C and 42^°^C). These data demonstrate that the RNA structure around the RBS is modulated by sequence variation, suggesting that the polymorphisms modulate initiation of protein synthesis.

In order to examine the impact of the polymorphisms on fHbp expression, we generated a series of isogenic ST-41/44 complex strains with combinations of the SNPs, *fHbp*_S-7_T/C and *fHbp*_S13_A/G, and examined surface expression of fHbp. Notably, the *fHbp*_S-7_T IMD risk allele conferred significantly higher fHbp expression, measured by flow cytometry, than *fHbp*_S-7_C, irrespective of *fHbp*_S13_A/G (*p*<0.05, **Figure 4D-E**). Further, when bacteria were incubated in normal human sera (NHS), strains with *fHbp*_S-7_T displayed increased survival compared with *fHbp*_S-7_C, but not in heat-inactivated human serum lacking functional complement, irrespective of the *fHbp*_S13_A/G allele (*p*<0.05, One way Anova, **Figure 4F, Supplementary Figure 15**). Taken together, these results are consistent with the IMD-associated alleles at the 5’ end of *fHbp* around the conferring enhanced resistance of bacteria against complement-mediated killing, a major component of immunity against *N. meningitidis*.

Our study exclusively used publicly available genome sequences and metadata stored in the pubMLST *Neisseria* database (https://pubmlst.org/neisseria/), using well-described datasets from the Czech Republic and of clonal complex ST-41/44 isolates for replication. GWAS studies of virulence are particularly suitable in recombinogenic organisms such as *N. meningitidis*, as recombination assists fine-mapping by breaking down clonal background [30, 31, 8, 10, 32]. Previous GWAS have not identified variants influencing IMD severity, including in *fHbp*, and adaptation has also not between identified between paired isolates sampled from blood and cerebrospinal fluid [33, 34]. We found that the genetic architecture of meningococcal virulence is polygenic, adding to the growing understanding on virulence factors influencing the risk of IMD [17, 18, 35, 36]. Although several loci across the genome were identified, the major signals associated with IMD *versus* carriage emanated from the *fba-fHbp* region. This is particularly significant as GWAS of human genetic susceptibility identified SNPs in CFH associated with IMD, although the mechanism remains unknown [4, 37]. Our results therefore highlight that the interaction of bacterial and human gene pools across a single molecular interface, involving fHbp and CFH, dictates host susceptibility and the propensity of strains to cause invasive disease.

## MATERIALS AND METHODS

### Sampling frames

The discovery sample collection comprised 261 *Neisseria meningitidis* isolates from the Czech Republic [8, 9, 10]. Carriage samples were collected from young adults with no association with patients with IMD over four months, while disease isolates were from cases of IMD submitted to the Czech National Reference Laboratory for Meningococcal Infections in 1993 [8, 10]. 252 isolates from this collection were described previously to identify the MDA island by PCR, but not by WGS. Illumina sequencing reads were downloaded from the European Nucleotide Archive (ENA, http://www.ebi.ac.uk/ena), and Velvet *de novo* assemblies from PubMLST (https://pubmlst.org/neisseria/) [24]. PubMLST IDs, ENA accession numbers and phenotypes can be found in **Supplementary Table 6**.

The replication sample collection comprised 1,295 ST-41/44 complex genomes downloaded from pubMLST. We downloaded all genomes with a non-empty disease or carriage status, with *de novo* assemblies ≥2Mb in length, excluding the 261 genomes from the discovery sample collection and excluding genomes that met any of the following criteria: i) annotated as non-*Neisseria meningitidis*, ii) annotated with the disease phenotype “other”, iii) non-ST-41/44 complex assignment (described below), iv) genomes with more than 700 contigs, v) genomes with only one or two contigs, and vi) genomes with a total assembly length greater than 2.5Mb. PubMLST IDs and phenotypes can be found in **Supplementary Table 7**.

### SNP calling and kmer counting

For the discovery sample collection, sequence reads were mapped against the reference ST-11 complex, FAM18 genome (GenBank number: NC_008767.1) using Stampy [38]. Bases were called using previously described quality filters [39, 40, 41]. We identified 150,502 biallelic SNPs, 6,063 tri-allelic SNPs, and 239 tetra-allelic SNPs.

To capture non-SNP-based variation, and SNPs not in the reference genome, we pursued a kmer-based approach where all unique 31 bp haplotypes were counted from Velvet assemblies using dsk [42] in both sample collections. For both sample collections, a unique set of variably present kmers across each data set was created, with the presence or absence of each unique kmer determined per genome. Algorithms, coded in C++, can be downloaded from https://github.com/jessiewu/bacterialGWAS/blob/master/kmerGWAS/gwas_kmer_pattern [16]. We identified 7,806,583 variably present kmers in the discovery sample collection and 11,114,868 in the replication study collection.

### Phylogenetic inference

A maximum likelihood phylogeny was estimated for the discovery study collection for visualisation of phylogenetic relationships in the sampling frame and for SNP imputation purposes using RaxML [43] with a general time reversible (GTR) model and no rate heterogeneity, using alignments from the mapped data based on biallelic sites, with non-biallelic sites being set to the reference allele.

Non-ST-41/44 complex assignment in the replication study collection was determined using the kmer counts. A UPGMA tree was built using a kmer presence/absence distance matrix and all descendants of the most recent common ancestor of the genomes annotated in pubMLST as ST-41/44 complex were kept in order to identify unlabelled members of the complex.

### SNP imputation

For the SNP discovery analysis, missing sites due to sequencing ambiguity or strict SNP-calling thresholds were imputed using ancestral state reconstruction [44] implemented in ClonalFrameML [45]. This approach was previously shown to achieve high accuracy [16].

### Correcting for multiple testing

Multiple testing was accounted for by applying Bonferroni correction to control the strong-sense family-wise error rate (ssFWER) [46]. The unit of correction for all studies of individual loci in the discovery GWAS was taken to be the number of unique “phylopatterns” *i*.*e*. the number of unique partitions of individuals according to allele membership. The locus effect of an individual variant was considered to be significant if its *P* value was smaller than *α*/*n*_p_, where we took *α* = 0.05 to be the genome-wide false positive rate (*i*.*e*. family-wide error rate, FWER) and *n*_p_ to be the number of unique phylopatterns. In the discovery SNP-based analysis, *n*_p_ was taken to be the number of unique SNP phylopatterns (80,099) and in the kmer-based analyses, *n*_p_ was taken to be the number of unique kmer presence/absence phylopatterns (307,830).

In the replication GWAS, since we tested whether the two genome-wide significant, disease-associated kmers in the *fba*-*fHbp* operon replicated, we applied a Bonferroni correction to obtain a significance threshold of 0.05/2=10^−1.60^.

When testing the discovery GWAS for lineage effects by the Wald test for principal component-phenotype associations, a Bonferroni correction was applied for the number of non-redundant principal components, which equalled the sample size (261) since no two genomes were identical.

### Testing for locus effects

We performed association testing using the R package *bugwas* (https://github.com/sgearle/bugwas) [16] which uses linear mixed model (LMM) analyses implemented in the software GEMMA [13] to control for population structure. We modified GEMMA version 0.93 to enable significance testing of non-biallelic variants [16]. GEMMA was run using no minor allele frequency cut-off to include all variants.

For the discovery GWAS, we computed the relatedness matrix from biallelic SNPs only using the option “-gk 1” in GEMMA to calculate the centred relatedness matrix. For the replication study, we computed the relatedness matrix from the kmer presence/absence matrix using Java code which also calculates the centred relatedness matrix.

### Identifying lineage effects

We tested for associations between bacterial lineages and the phenotype in the discovery sample collection using the R package *bugwas* (https://github.com/sgearle/bugwas) [16]. Principal components were computed based on biallelic SNPs and interpreted in terms of bacterial lineages. To test the null hypothesis of no background effect of each principal component we employed a Wald test, where we compared the test statistic against a χ^2^ distribution with one degree of freedom to obtain a *P* value.

### Estimating sample heritability

Heritability of the sample, the proportion of the phenotypic variation that can be explained by the bacterial genotype, was estimated using the LMM null model in GEMMA from post-imputation biallelic SNPs [13]. Estimating heritability in case control studies is dependent on the prevalence of cases and the sampling scheme [47]. The proportion of sample heritability explained by the kmers *fba*_K898_ and *fba*_K899_ in the discovery set was estimated by including the phylopattern of the two kmers as a covariate in the LMM null model in GEMMA, and calculating the difference in heritability compared to including no covariates.

### Testing for independent SNP associations

To determine whether pairs of significant SNP associations in the discovery sample collection represented independent signals, the two unique significant SNP patterns were tested using LMM including both SNPs as fixed effects, thereby assuming additivity between the two loci.

### Variant annotation

SNPs were annotated in R using scripts at http://github.com/jessiewu/bacterialGWAS. The reference FASTA and GenBank files were used in order to determine SNP type (synonymous, non-synonymous, nonsense, read-through and intergenic), codon, codon position, reference and non-reference amino acids, gene name and gene product, on the assumption of a single change to the reference sequence.

To annotate kmer sequences, we mapped kmers to the reference FAM18 genome using Bowtie2 [48] and the options “-r -D 24 -R 3 -N 0 -L 18 -i S,1,0.30” to identify a single best mapping position for each kmer. For kmers which did not map to the reference genome, BLAST [49] was used to identify the kmer position within FAM18. BLAST results of any sequence length were taken, and the number of mismatches along the whole length of the kmer was recalculated assuming the whole kmer aligned. Kmers with five or fewer mismatches to the reference were shown as aligned to the reference, all other kmers were shown as unaligned to the reference.

To understand the variation captured by the significant kmers in the gene *csb*, BLAST [49] was used to extract all copies of the MC58 (Genbank accession number NC_003112.2) allele of *csb*, the allele that the significant *csb* kmers mapped to.

As the reference FAM18 genome contains multiple copies of the gene *tspB*, to understand the variation captured by the significant kmers in *tspB*, BLAST [49] was used to identify all kmer alignments with just the FAM18 *tspB* gene NMC_RS00140.

### Software

Software applied within these analyses can be found at http://github.com/jessiewu/bacterialGWAS and http://github.com/sgearle/bugwas.

### Strain construction

The primers and strains used to test the effects of SNPs are listed in **Supplementary Tables 2-4**. The *fba*_S897_/_S900_ SNPs were constructed by inserting a Kanamycin resistance cassette downstream of *fHbp*. First, the upstream fragment (starting 843 bp upstream of the *fHbp* start codon including the C terminus of *fba*, terminating 12 bp downstream of the *fHbp* stop codon) and downstream fragment (751 bp downstream of the *fHbp* stop codon) were amplified with primers ERS001/ERS004 and ERS007/ERS008 respectively from 0011/93 *N. meningitidis* gDNA. The kanamycin resistance cassette was amplified from pGEMTEasy-Kan using ERS005/ERS006 and the three fragments were cloned into pUC19 using NEB Builder HiFi DNA assembly kit (New England Biolabs). A second set of overlap primers were used to introduce SNPs into a second upstream fragment using primer combinations: ERS001/ERS002 and ERS003/ERS004, ERS001/ERS009 and ERS010/ERS004, and ERS001/ERS011 and ERS012/ERS004. The constructs were purified and transformed into 0011/93 *N. meningitidis*. For each strain, three independent single colonies were pooled and gDNA from the pooled stocks was checked by PCR and sequencing.

The *fHbp*_S-7_/_S13_ SNPs were constructed by inserting an erythromycin resistance cassette downstream of *fHbp*. First, a fragment corresponding to 496 bp upstream of the *fHbp* start codon and the *fHbp* ORF, and a fragment corresponding to 707 bp downstream of the *fHbp* stop codon) were amplified with primers ML428/ML429 and ML434/ML433 respectively from 0011/93 *N. meningitidis* gDNA. The erythromycin resistance cassette was amplified from pNMC2 [50] using ML430/ML435 and the three fragments were cloned into pUC19 using NEB Builder HiFi DNA assembly kit (New England Biolabs). The resulting vector was used as a template to generate *fHbp* with different SNPs by site directed mutagenesis using primer combinations: ML436/405 and ML437/406, ML438/ML405 and ML439/ML406, and ML440/ML405 and ML441/406. The constructs were purified and used to transform 0011/93 *N. meningitidis*. For each strain, three independent single transformants were pooled and gDNA from the pooled stocks was checked by PCR and sequencing.

### Generation of plasmids and protein purification

V2.24 *fHbp* was amplified from *N. meningitidis* OX99.32412 and SNPs introduced by PCR, then ligated into pET21b using Quick-Stick Ligase (Bioline). Versions of *fHbp* were ligated into pET28a-His-MBP-TEV (in frame with sequence encoding a histidine tag and the *Escherichia coli* maltose-binding protein (MBP) with a C-terminal TEV cleavage site) linearised with *Xho*I, and constructs confirmed by sequencing.

v2.24 fHbps were expressed in *E. coli* B834 during growth at 22°C for 24 hrs with 1 mM IPTG (final concentration). Bacteria were harvested and resuspended in Buffer A (50 mM Na-phosphate pH 8.0, 300 mM NaCl, 30 mM imidazole) and the fHbp purified by Nickel affinity chromatography (Chelating Sepharose Fast Flow; GE Healthcare). Columns were washed with Buffer A, then with 80:20 Buffer A:Buffer B (50 mM Na-phosphate pH 8.0, 300 mM NaCl, 300 mM Imidazole), and proteins eluted in 40:60 Buffer A:Buffer B. Proteins were dialysed overnight at 4^°^C into PBS, 1mM DTT pH 8.0 with TEV protease prior to Nickel affinity chromatography to remove the HIS-GST-TEV. fHbp was eluted from Sepharose columns with Buffer B after washing with buffer C (50 mM Na-phosphate pH 6.0, 500 mM NaCl, 30 mM Imidazole), and dialysed overnight at 4^°^C into Tris pH 8.0. fHbp v1.1^I311A^ expression and purification was performed as described previously [51].

### Electrophoresis mobility shift assays

Gel retardation assays were carried out as previously using purified FNR^D154A^, which forms functional FNR dimers under aerobic conditions [52]. Sequences upstream of *fHbp* were amplified with primers ERS012/013, and the full length (420 bp) or *Hae*III-digested (294 and 126 bp) fragments end-labelled with [γ-^32^P]-ATP with T4 polynucleotide kinase (New England BioLabs). Approximately 0.5 ng of each labelled fragment was incubated with varying amounts of FNR^D154A^ in 10 mM potassium phosphate (pH 7.5), 100 mM potassium glutamate, 1 mM EDTA, 50 μM DTT, 5% glycerol and herring sperm DNA (25 μg ml^−1^). After incubation at 37°C for 20 min, samples were separated on 6% polyacrylamide gels containing 2% glycerol. Gels were analysed using a Bio-Rad Molecular Imager FX and Quantity One software (Bio-Rad).

### CFH binding ELISA

To investigate CFH binding by ELISA, 96-well plates (F96 MaxiSorp; Nunc) were coated with recombinant fHbp (5 μg/well) overnight at 4^°^C prior to blocking with 3% bovine serum albumin (BSA) in PBS with 0.05% Tween 20 at 37^°^C. Plates were incubated with dilutions of CFH (Sigma). Binding was detected with anti-CFH mAb (OX24) and HRP-conjugated goat anti-mouse polyclonal antibody (Dako), and visualized with 3,3’,5,5’-tetramethylbenzidine (TMB) substrate reagent (Roche) and 2 N sulphuric acid stop solution (Roche) according to the manufacturer’s instructions, and the A_450_ measured (SpectraMax M5; Molecular Devices).

### Serum assays

Pooled normal human serum (NHS) were used in serum assays, and heat inactivated (NHS-HI) by heating at 56°C for 30 min. *N. meningitidis* was grown overnight on Brain Heart Infusion (BHI) agar, and then 10^4^ CFU were incubated in dilutions of NHS or NHS-HI for 30 min at 37°C in the presence of CO_2_. Bacterial survival was determined by plating onto BHI agar in triplicate. Percent survival was calculated by comparing bacterial recovery in serum with recovery from samples containing no serum. Significance was analysed by two-way ANOVA (GraphPad Prism v.8.0).

### Flow cytometry

*N. meningitidis* was grown on BHI agar at 32°C or 37°C prior to fixation for two hours in 3% paraformaldehyde. Surface localisation of fHbp was detected using anti-fHbp pAbs and goat anti-mouse IgG-Alexa Fluor 647 conjugate (Molecular Probes, LifeTechnologies). Samples were run on a FACSCalibur (BD Biosciences), and at least 10^4^ events recorded before results were analysed by calculating the geometric mean fluorescence intensity in FlowJo vX software (Tree Star).

### SHAPE RNA secondary structure analysis

SHAPE experiments were performed using RNA transcribed *in vitro* from cDNA sequence [53]. The DNA templates contained a double-stranded T7 RNA polymerase promoter sequence (TTCTAATACGACTCACTATA) followed by the sequence of interest (**Supplementary Table 5**). RNA purification was done with an RNA clean kit (Zymo research); RNA concentrations were measured on a Nanodrop 100 spectrophotometer. RNA chemical modification was performed in volumes of 30µl with 1.5pmol of RNA within Folding buffer (50 mM HEPES pH 8.0, 16.5 mM MgCl_2_). RNA samples were pre-heated at 65°C for 3 mins and immediately incubated at 30°C, 37°C or 42°C water baths for 30 mins. The modification reagent N-methylisatoic anhydride (NMIA) was added at increasing concentrations between 0 and 13mM, with DMSO (no NMIA) as control. Modification reactions were incubated for another 45mins before ethanol precipitation [54, 55]. Reverse transcription was performed using Super Script III reverse transcriptase (Invitrogen). ^32^P-labeled reverse transcription primers (GV1-3) are listed in **Supplementary Table 4**. Electrophoresis on 8% (vol/vol) polyacrylamide gels was then performed to separate fragments. Band-intensities were quantified using SAFA, version 1.1 Semi-Automated Footprinting Analysis [56].

All structure calculations were performed using RNAstructure software [57]. ΔG°SHAPE free energy change values were added to the thermodynamic free energy parameters for each nucleotide [58, 59]. Pseudoknot-energy parameters were used in calculation of ΔG°(SHAPE), according to the equation, Δ*G*°*SHAPE(i)= m ln[SHAPE reactivity(i)+ 1]+ b*. In this analysis, parameters were optimized at m=0.3 and b=-1.2 kcal/mol for fHbps7T/s13A; m=0.4 and b=-2.0 kcal/mol for fHbps-7C/s13G; nucleotides with normalized SHAPE reactivities 0–0.40, 0.40– 0.85, and >0.85 correspond to low, medium, and highly reactive positions, respectively [58, 59]. Secondary structures were rendered using VARNA [60]. [1]

### Harmonic mean *P* value

The harmonic mean *p*-value (HMP) method performs a combined test of the null hypothesis that no *p*-value is significant [61]. The HMP method controls for the ssFWER like the Bonferroni correction. We applied the HMP procedure to the *fba*-*fHbp* region in the discovery and replication studies, including all unique kmer phylopatterns that mapped to either of the two genes plus their upstream intergenic regions. We calculated the asymptotically exact *p*-values using the p.hmp function from the R package ‘harmonicmeanp’, giving equal weight to all kmer phylopatterns, and the total number of tests performed genome-wide was set to be the number of kmer phylopatterns (*n*_p_) in order to control the genome-wide ssFWER despite analysing just the *fba*-*fHbp* region. We adjusted the *p*-value by dividing it by the sum of the weights of the kmer phylopatterns included in the *fba*-*fHbp* region so that it could be directly compared to alpha, the intended ssFWER, which we set to be 0.05.

## Data Availability

Illumina sequencing reads were downloaded from the European Nucleotide Archive (ENA, http://www.ebi.ac.uk/ena), and Velvet *de novo* assemblies and phenotypes from PubMLST (https://pubmlst.org/neisseria/). PubMLST IDs, ENA accession numbers and phenotypes can be found in Supplementary Tables 6 and 7.

## Data Availability

All data, code, and materials used in the analyses can be accessed at:https://github.com/SiyuChenOxf/COVID19SeroModel/tree/master

## ACKNOWLEDGEMENTS

Work in CMT’s laboratory is funded by a Wellcome Trust Senior Investigator award. D.J.W. is supported by a Sir Henry Dale Fellowship (Grant 101237/Z/13/B) and a Big Data Institute Robertson Fellowship. Computation using the Oxford Biomedical Research Computing (BMRC) facility is supported by Health Data Research UK and the NIHR Oxford Biomedical Research Centre. Financial support was provided by the Wellcome Trust Core Award Grant Number 203141/Z/16/Z. Work at the University of Washington was supported by NIH NIGMS 1R35 GM 126942.

## REFERENCES

[1] B. Wang, R. Santoreneos, L. Giles, H. H. A. Afzali and H. Marshall, “Case fatality rates of invasive meningococcal disease by serogroup and age: A systematic review and meta-analysis,” Vaccine, vol. 37, no. 21, pp. 2768–2782, 2019.

[2] H. Christensen, M. May, L. Bowen, M. Hickman and C. L. Trotter, “Meningococcal carriage by age: a systematic review and meta-analysis,” The Lancet Infectious Diseases, vol. 10, no. 12, pp. 853–861, 2010.

[3] B. P. Morgan and M. J. Walport, “Complement deficiency and disease,” Immunology Today, vol. 12, no. 9, pp. 301–306, 1991.

[4] S. Davila, V. J. Wright, C. C. Khor, K. S. Sim, A. Binder, W. B. Breunis, D. Inwald, S. Nadel, H. Betts, E. D. Carrol, R. de Groot, P. W. Hermans, J. Hazelzet, M. Emonts, C. C. Lim, T. W. Kuijpers, F. Martinon-Torres, A. Salas, W. Zenz, M. Levin and M. L. Hibberd, “Genome-wide association study identifies variants in the CFH region associated with host susceptibility to meningococcal disease,” Nature Genetics, vol. 42, no. 9, pp. 772– 776, 2010.

[5] A. Biebl, A. Muendlein, E. Kinz, H. Drexel, M. Kabesch, W. Zenz, R. Elling, C. Müller, T. Keil, S. Lau and B. Simma, “Confirmation of Host Genetic Determinants in the CFH Region and Susceptibility to Meningococcal Disease in a Central European Study Sample.,” The Pediatric infectious disease journal, vol. 34, no. 10, pp. 1115–1117, 2015.

[6] M. C. Schneider, R. M. Exley, H. Chan, I. Feavers, Y.-H. Kang, R. B. Sim and C. M. Tang, “Functional Significance of Factor H Binding to Neisseria meningitidis,” The Journal of Immunology, vol. 176, no. 12, pp. 7566–7575, 2006.

[7] G. Madico, J. A. Welsch, L. A. Lewis, A. McNaughton, D. H. Perlman, C. E. Costello, J. Ngampasutadol, U. Vogel, D. M. Granoff and S. Ram, “The Meningococcal Vaccine Candidate GNA1870 Binds the Complement Regulatory Protein Factor H and Enhances Serum Resistance,” The Journal of Immunology, vol. 177, no. 1, pp. 501–510, 2006.

[8] K. A. Jolley, J. Kalmusova, E. J. Feil, S. Gupta, M. Musilek, P. Kriz and M. C. J. Maiden, “Carried meningococci in the Czech Republic: A diverse recombining population,” Journal of Clinical Microbiology, vol. 38, no. 12, pp. 4492–4498, 2000.

[9] K. A. Jolley, J. Kalmusova, E. J. Feil, S. Gupta, M. Musilek, P. Kriz and M. C. J. Maiden, “Carried Meningococci in the Czech Republic: a Diverse Recombining Population,” Journal of Clinical Microbiology, vol. 40, no. 9, pp. 3549–3550, 2002.

[10] K. A. Jolley, D. J. Wilson, P. Kriz, G. Mcvean and M. C. J. Maiden, “The Influence of Mutation, Recombination, Population History, and Selection on Patterns of Genetic Diversity in Neisseria meningitidis,” Molecular Biology and Evolution, vol. 22, no. 3, pp. 562–569, 2005.

[11] M. C. J. Maiden, J. A. Bygraves, E. Feil, M. G. J. E. Russell, R. Urwin, Q. Zhang, J. Zhou, K. Zurth, D. A. Caugant, I. M. Feavers, M. Achtman and B. G. Spratt, “Multilocus sequence typing: A portable approach to the identification of clones within populations of pathogenic?microorganisms,” Proceedings of the National Academy of Sciences of the United States of America, vol. 95, no. 6, pp. 3140–3145, 1998.

[12] S. Budroni, E. Siena, J. C. Dunning Hotopp, K. L. Seib, D. Serruto, C. Nofroni, M. Comanducci, D. R. Riley, S. C. Daugherty, S. V. Angiuoli, A. Covacci, M. Pizza, R. Rappuoli, E. R. Moxon, H. Tettelin and D. Medini, “Neisseria meningitidis is structured in clades associated with restriction modification systems that modulate homologous recombination.,” PNAS, vol. 108, no. 11, pp. 4494–4499, 2011.

[13] X. Zhou and M. Stephens, “Genome-wide efficient mixed-model analysis for association studies,” Nature Genetics, vol. 44, no. 7, pp. 821–824, 2012.

[14] S. D. Bentley, G. S. Vernikos, L. A. S. Snyder, C. Churcher, C. Arrowsmith, T. Chillingworth, A. Cronin, P. H. Davis, N. E. Holroyd, K. Jagels, M. Maddison, S. Moule, E. Rabbinowitsch, S. Sharp, L. Unwin, S. Whitehead, M. A. Quail, M. Achtman, B. Barrell, N. J. Saunders and J. Parkhill, “Meningococcal genetic variation mechanisms viewed through comparative analysis of serogroup C strain FAM18,” PLoS Genetics, vol. 3, no. 2, pp. 0230–0240, 2007.

[15] S. K. Sheppard, X. Didelot, G. Meric, A. Torralbo, K. a. Jolley, D. J. Kelly, S. D. Bentley, M. C. J. Maiden, J. Parkhill and D. Falush, “Genome-wide association study identifies vitamin B5 biosynthesis as a host specificity factor in Campylobacter,” Proceedings of the National Academy of Sciences of the United States of America, vol. 110, no. 29, pp. 11923–11927, 2013.

[16] S. G. Earle, C.-h. Wu, J. Charlesworth, N. Stoesser, N. C. Gordon, T. M. Walker, C. C. A. Spencer, Z. Iqbal, D. A. Clifton, K. L. Hopkins, N. Woodford, E. G. Smith, N. Ismail, M. J. Llewelyn, T. E. Peto, D. W. Crook, G. McVean, A. S. Walker and D. J. Wilson, “Identifying lineage effects when controlling for population structure improves power in bacterial association studies,” Nature Microbiology, vol. 1, no. 5, p. 16041, 2016.

[17] E. Bille, J.-R. Zahar, A. Perrin, S. Morelle, P. Kriz, K. Jolley, M. C. J. Maiden, C. Dervin, X. Nassif and C. R. Tinsley, “A chromosomally integrated bacteriophage in invasive meningococci.,” J Exp Med, vol. 201, no. 12, pp. 1905–1913, 2005.

[18] E. Bille, R. Ure, S. J. Gray, E. B. Kaczmarski, N. D. McCarthy, X. Nassif, M. C. J. Maiden and C. R. Tinsley, “Association of a bacteriophage with meningococcal disease in young adults.,” PloS one, vol. 3, no. 12, p. e3885, 2008.

[19] M. G. Müller, J. Y. Ing, M. K.-W. Cheng, B. A. Flitter and G. R. Moe, “Identification of a Phage-Encoded Ig-Binding Protein from Invasive Neisseria meningitidis,” The Journal of Immunology, vol. 191, no. 6, pp. 3287–3296, 2013.

[20] M. G. Müller, N. E. Moe, P. Q. Richards and G. R. Moe, “Resistance of Neisseria meningitidis to Human Serum Depends on T and B Cell Stimulating Protein B,” Infection and Immunity, vol. 83, no. 4, pp. 1257–1264, 2015.

[21] M. V. R. Weber, H. Claus, M. C. J. Maiden, M. Frosch and U. Vogel, “Genetic mechanisms for loss of encapsulation in polysialyltransferase-gene-positive meningococci isolated from healthy carriers,” International Journal of Medical Microbiology, vol. 296, no. 7, pp. 475–484, 2006.

[22] S. A. Tunio, N. J. Oldfield, A. Berry, D. A. A. Ala’Aldeen, K. G. Wooldridge and D. P. J. Turner, “The moonlighting protein fructose-1, 6-bisphosphate aldolase of Neisseria meningitidis: Surface localization and role in host cell adhesion,” Molecular Microbiology, vol. 76, no. 3, pp. 605–615, 2010.

[23] F. Shams, N. J. Oldfield, S. K. Lai, S. A. Tunio, K. G. Wooldridge and D. P. J. Turner, “Fructose-1,6-bisphosphate aldolase of Neisseria meningitidis binds human plasminogen via its C-terminal lysine residue,” MicrobiologyOpen, vol. 5, no. 2, pp. 340–350, 2016.

[24] K. Jolley, J. Bray and M. Maiden, “Open-access bacterial population genomics: BIGSdb software, the PubMLST.org website and their applications [version 1; peer review: 2 approved],” Wellcome Open Research, vol. 3, no. 124, 2018.

[25] H. B. Bratcher, C. Brehony, S. Heuberger, D. Pieridou-Bagatzouni, P. Křížová, S. Hoffmann, M. Toropainen, M. K. Taha, H. Claus, G. Tzanakaki, T. Erdôsi, J. Galajeva, A. van der Ende, A. Skoczyńska, M. Pana, A. Vaculíková, M. Paragi and Maide, “Establishment of the European meningococcal strain collection genome library (EMSC-GL) for the 2011 to 2012 epidemiological year,” Euro Surveill, vol. 23, no. 20, pp. 17–00474, 2018.

[26] D. G. Clayton, N. M. Walker, D. J. Smyth, R. Pask, J. D. Cooper, L. M. Maier, L. J. Smink, A. C. Lam, N. R. Ovington, H. E. Stevens, S. Nutland, J. M. M. Howson, M. Faham, M. Moorhead, H. B. Jones, M. Falkowski, P. Hardenbol, T. D. Willis and J. A. Todd, “Population structure, differential bias and genomic control in a large-scale, casecontrol association study,” Nature Genetics, vol. 37, no. 11, pp. 1243–1246, 2005.

[27] B. F. Voight and J. K. Pritchard, “Confounding from Cryptic Relatedness in Case-Control Association Studies,” PLOS Genetics, vol. 1, no. 3, p. e32, 2005.

[28] F. Oriente, V. Scarlato and I. Delany, “Expression of factor H binding protein of meningococcus responds to oxygen limitation through a dedicated FNR-regulated promoter,” Journal of Bacteriology, vol. 192, no. 3, pp. 691–701, 2010.

[29] E. Loh, H. Lavender, F. Tan, A. Tracy and C. Tang, “Thermoregulation of Meningococcal fHbp, an Important Virulence Factor and Vaccine Antigen, Is Mediated by Anti-ribosomal Binding Site Sequences in the Open Reading Frame,” PLoS Pathogens, vol. 12, no. 8, p. e1005794, 2016.

[30] E. J. Feil, M. C. Maiden, M. Achtman and B. G. Spratt, “The relative contributions of recombination and mutation to the divergence of clones of Neisseria meningitidis.,” Molecular Biology and Evolution, vol. 16, no. 11, pp. 1496–1502, 1999.

[31] E. C. Holmes, M. C. Maiden and R. Urwin, “The influence of recombination on the population structure and evolution of the human pathogen Neisseria meningitidis.,” Molecular Biology and Evolution, vol. 16, no. 6, pp. 741–749, 1999.

[32] D. Falush and R. Bowden, “Genome-wide association mapping in bacteria?,” Trends in microbiology, vol. 14, no. 8, pp. 353–355, 2006.

[33] P. H. C. Kremer, J. A. Lees, B. Ferwerda, A. van de Ende, M. C. Brouwer, S. D. Bentley and D. van de Beek, “Genetic Variation in Neisseria meningitidis Does Not Influence Disease Severity in Meningococcal Meningitis,” Frontiers in Medicine, vol. 7, p. 826, 2020.

[34] J. Lees, P. Kremer, A. Manso, N. Croucher, B. Ferwerda, M. Serón, M. Valls Oggioni, J. Parkhill, M. Brouwer, A. Ende, D. Beek and S. Bentley, “Large scale genomic analysis shows no evidence for pathogen adaptation between the blood and cerebrospinal fluid niches during bacterial meningitis,” Microbial Genomics, vol. 3, no. 1, p. e000103, 2017.

[35] J. C. Dunning Hotopp, R. Grifantini, N. Kumar, Y. L. Tzeng, D. Fouts, E. Frigimelica, M. Draghi, M. M. Giuliani, R. Rappuoli, D. S. Stephens, G. Grandi and H. Tettelin, “Comparative genomics of Neisseria meningitidis: Core genome, islands of horizontal transfer and pathogen-specific genes,” Microbiology, vol. 152, no. 12, pp. 3733–3749, 2006.

[36] C. Collins and X. Didelot, “A phylogenetic method to perform genome-wide association studies in microbes that accounts for population structure and recombination,” PLOS Computational Biology, vol. 14, no. 2, p. e1005958, 2018.

[37] F. Martinón-Torres, E. Png, C. Khor, S. Davila, V. Wright, K. Sim, A. Vega,,. Fachal, D. Inwald, S. Nadel, E. Carrol, N. Martinón-Torres, S. Alonso, A. Carracedo, E. Morteruel and J. López-Bayón, “Natural resistance to Meningococcal Disease related to CFH loci: Meta-analysis of genome-wide association studies,” Scientific Reports, vol. 6, no. 1, p. 35842, 2016.

[38] G. Lunter and M. Goodson, “Stampy: a statistical algorithm for sensitive and fast mapping of Illumina sequence reads.,” Genome research, vol. 21, no. 6, pp. 936–939, 2011.

[39] X. Didelot, R. Bowden, D. J. Wilson, T. E. A. Peto and D. W. Crook, “Transforming clinical microbiology with bacterial genome sequencing,” Nature Reviews Genetics, vol. 13, no. 9, pp. 601–612, 2012.

[40] B. C. Young, T. Golubchik, E. M. Batty, R. Fung, H. Larner-svensson, A. J. Rimmer, M. Cule, C. L. C. Ip, X. Didelot, R. M. Harding, P. Donnelly, T. E. Peto, D. W. Crook, R. Bowden and D. J. Wilson, “Evolutionary dynamics of Staphylococcus aureus during progression from carriage to disease,” Proceedings of the National Academy of Sciences of the United States of America, vol. 109, no. 12, pp. 4550–4555, 2012.

[41] T. Golubchik, E. Batty, R. Miller, H. Farr, B. Young, H. Larner-Svensson, R. Fung, H. Godwin, K. Knox, A. Votintseva, R. Everitt, T. Street, M. Cule, C. Ip, X. Didelot, T. Peto, R. Harding, D. Wilson, D. Crook and R. Bowden, “Within-host evolution of Staphylococcus aureus during asymptomatic carriage,” PloS One, vol. 8, no. 5, p. e61319, 2013.

[42] G. Rizk, D. Lavenier and R. Chikhi, “DSK: k-mer counting with very low memory usage,” Bioinformatics, vol. 29, no. 5, pp. 652–653, 2013.

[43] A. Stamatakis, “RAxML version 8: A tool for phylogenetic analysis and post-analysis of large phylogenies,” Bioinformatics, vol. 30, no. 9, pp. 1312–1313, 2014.

[44] T. Pupko, I. Pe, R. Shamir and D. Graur, “A Fast Algorithm for Joint Reconstruction of Ancestral Amino Acid Sequences,” Molecular Biology and Evolution, vol. 17, no. 6, pp. 890–896, 2000.

[45] X. Didelot and D. J. Wilson, “ClonalFrameML: Efficient Inference of Recombination in Whole Bacterial Genomes,” PLOS Computational Biology, vol. 11, p. e1004041, 2015.

[46] O. J. Dunn, “Estimation of the Medians for Dependent Variables,” The Annals of Mathematical Statistics, vol. 30, pp. 192-197, 1959.

[47] N. Zaitlen and P. Kraft, “Heritability in the genome-wide association era,” Human Genetics, vol. 131, no. 10, pp. 1655–1664, 2012.

[48] B. Langmead and S. L. Salzberg, “Fast gapped-read alignment with Bowtie 2,” Nature Methods, vol. 9, no. 4, pp. 357–359, 2012.

[49] C. Camacho, G. Coulouris, V. Avagyan, N. Ma, J. Papadopoulos, K. Bealer and T. L. Madden, “BLAST+: architecture and applications,” BMC Bioinformatics, vol. 10, no. 1, p. 421, 2009.

[50] M. Lobanovska, C. M. Tang and R. M. Exley, “Contribution of σ70 and σN Factors to Expression of Class II pilE in Neisseria meningitidis,” Journal of Bacteriology, vol. 201, no. 20, pp. e00170–19, 2019.

[51] S. Johnson, L. Tan, S. van der Veen, J. Caesar, E. Goicoechea De Jorge, R. J. Harding, X. Bai, R. M. Exley, P. N. Ward, N. Ruivo, K. Trivedi, E. Cumber, R. Jones, L. Newham and D. Staunton, “Design and Evaluation of Meningococcal Vaccines through Structure-Based Modification of Host and Pathogen Molecules,” PLOS Pathogens, vol. 8, no. 10, p. e1002981, 2012.

[52] D. Browning, C. Beatty, A. Wolfe, J. Cole and S. Busby, “Independent regulation of the divergent Escherichia coli nrfA and acsP1 promoters by a nucleoprotein assembly at a shared regulatory region,” Molecular Microbiology, vol. 43, no. 3, pp. 687–701, 2002.

[53] K. Weeks and D. Mauger, “Exploring RNA structural codes with SHAPE chemistry,” Accounts of Chemical Research, vol. 44, no. 12, pp. 1280–1291, 2011.

[54] J. B. Lucks, S. A. Mortimer, C. Trapnell, S. Luo, S. Aviran, G. P. Schroth, L. Pachter, J. A. Doudna and A. P. Arkin, “Multiplexed RNA structure characterization with selective 2’- hydroxyl acylation analyzed by primer extension sequencing (SHAPE-Seq),” PNAS, vol. 108, no. 27, pp. 11063–11068, 2011.

[55] L. Poulsen, L. Kielpinski, S. Salama, A. Krogh and J. Vinther, “SHAPE Selection (SHAPES) enrich for RNA structure signal in SHAPE sequencing-based probing data,” RNA, vol. 21, no. 5, pp. 1042–1052, 2015.

[56] R. Das, A. Laederach, S. Pearlman, D. Herschlag and R. Altman, “SAFA: semi-automated footprinting analysis software for high-throughput quantification of nucleic acid footprinting experiments,” RNA, vol. 11, no. 3, pp. 344–354, 2005.

[57] D. H. Mathews, M. D. Disney, J. L. Childs, S. J. Schroeder, M. Zuker and D. H. Turner, “Incorporating chemical modification constraints into a dynamic programming algorithm for prediction of RNA secondary structure,” PNAS, vol. 101, no. 19, pp. 7287– 7292, 2004.

[58] C. E. Hajdin, S. Bellaousov, W. Huggins, C. W. Leonard, D. H. Mathews and K. M. Weeks, “Accurate SHAPE-directed RNA secondary structure modeling, including pseudoknots,” PNAS, vol. 110, no. 14, pp. 5498–5503, 2013.

[59] K. Deigan, T. Li, D. Mathews and W. KM, “Accurate SHAPE-directed RNA structure determination,” PNAS, vol. 106, no. 1, pp. 91–102, 2009.

[60] K. Darty, A. Denise and Y. Ponty, “VARNA: Interactive drawing and editing of the RNA secondary structure,” Bioinformatics, vol. 25, no. 15, p. 1974, 2009.

[61] D. J. Wilson, “The harmonic mean p-value for combining dependent tests,” PNAS, vol. 116, no. 4, pp. 1195–1200, 2019.

