## Supplementary Figures, Tables and Text for "Polymorphisms affecting expression of the vaccine antigen factor H binding protein influence invasiveness of Neisseria meningitidis"

### SUPPLEMENTARY INFORMATION

**SUPPLEMENTARY FIGURES**

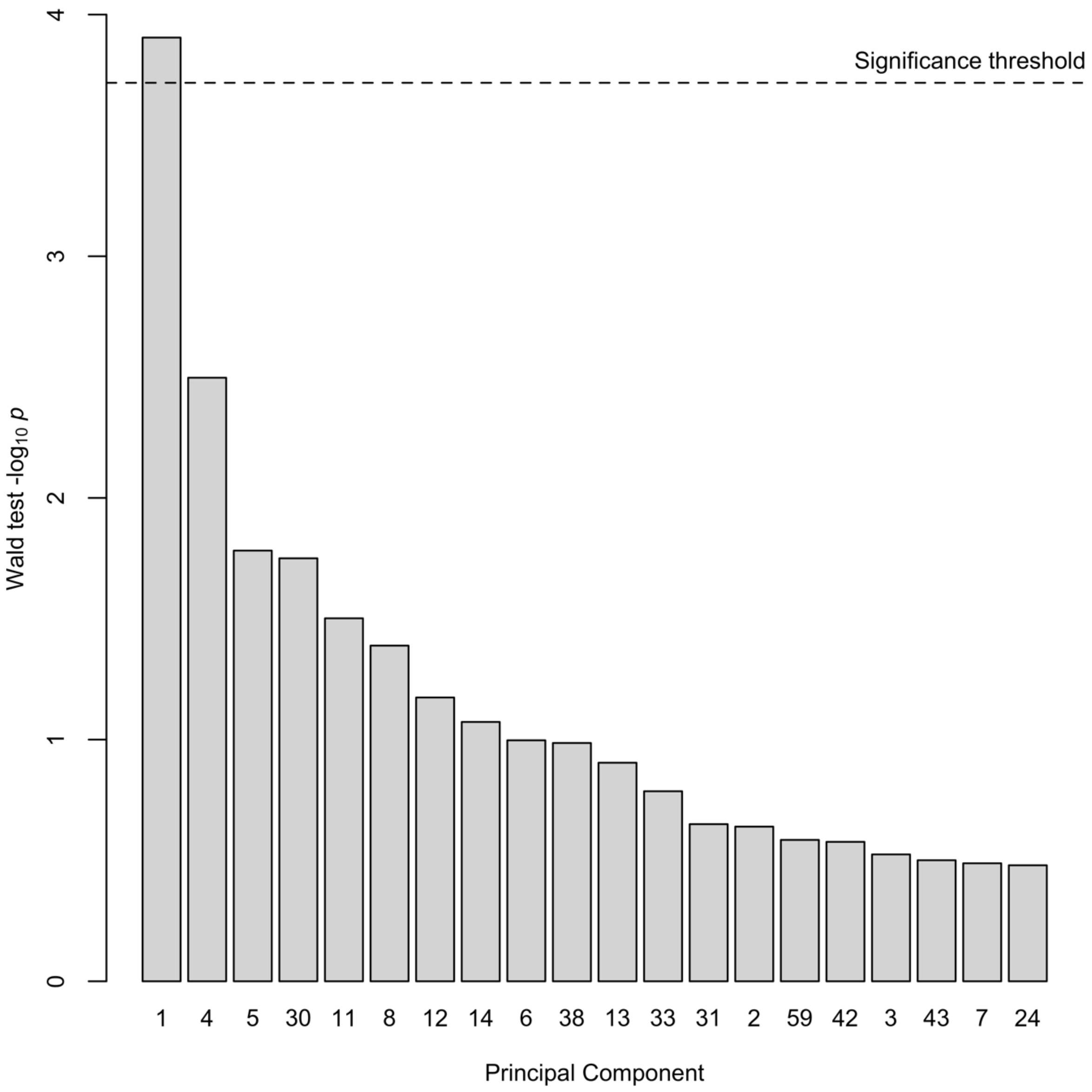


**Supplementary Figure 1** The 20 most significant principal components (PCs) by a Wald test, testing for association between PCs and case-control status. A Bonferroni correction was applied to the significance threshold for the number of non-redundant PCs.

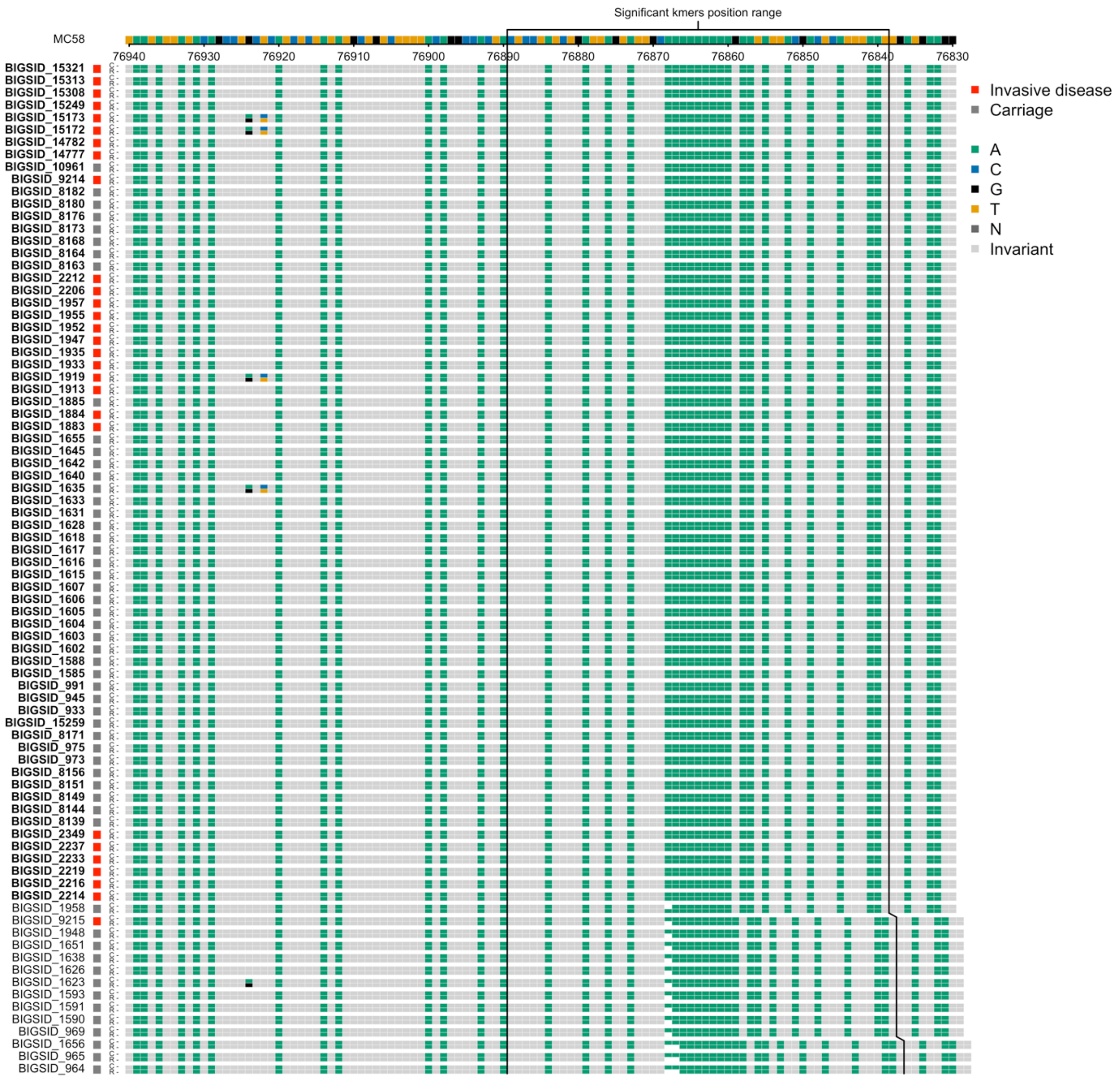


**Supplementary Figure 2** Alignments of the 82 genomes containing the MC58 (NC_003112.2) serogroup B *csb* gene in the discovery sample collection aligned with the MC58 *csb* gene positions 76940-76830 on the reverse strand, the coding strand for *csb* with all A alleles coloured. Each row shows an alignment of a contig (C) with the reference (R) from BLAST (Camacho et al. 2009). On the left, red indicates that the isolate was sampled from a patient with invasive disease, grey from a carrier. In the alignments, grey indicates identity between the contig and the reference. Polymorphisms are coloured by the alleles (A = green; C = blue; G = black; T = Orange) plus all invariant positions with the A allele are coloured green.

Insertions and deletions are shown in white. The top line shows the bases of the MC58 *csb* gene in the region. Vertical lines show the region where the 21 significant *csb* kmers mapped. Sample names are shown as BIGS IDs from pubMLST and sample names shown in bold contained the 21 significant kmers.

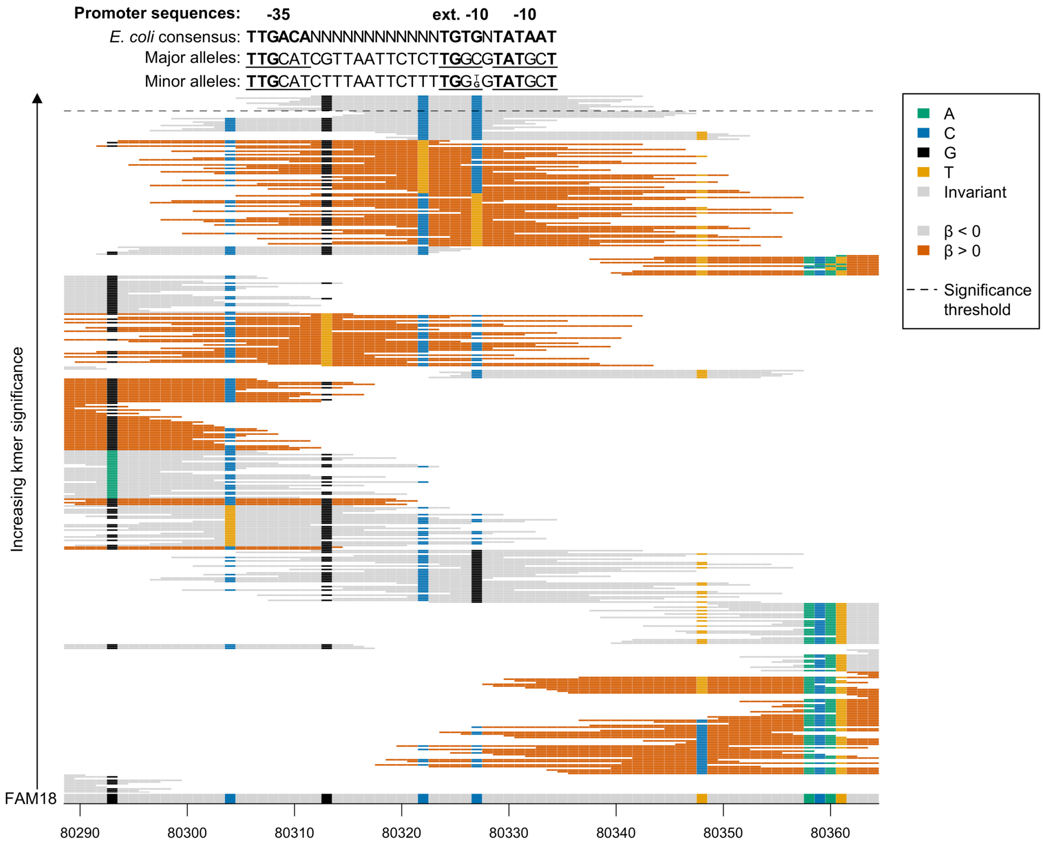


**Supplementary Figure 3** Close up of the significant kmers aligned to the intergenic region between *ctrE* and *ctrF* in the discovery sample collection. The reference genome FAM18 is shown at the bottom of the figure, grey for invariant sites and coloured at variant site positions. The kmers which map to the region shown are then plotted from least significant at the bottom to most significant at the top. The black dashed line indicates the Bonferroni-corrected significance threshold – all kmers above the line are significantly associated with the phenotype. The background colour of the kmers represents the direction of the association, grey when $\beta < 0$ (carriage-associated) and dark orange when $\beta > 0$ (disease-associated). Kmers are coloured by their allele at all variant positions (A = green; C = blue; G = black; T = Orange). The *E. coli* consensus for the -10 and -35 promoter regions are shown above the kmers aligned with the major and minor alleles in the discovery sample collection at these positions. Matches to the consensus are shown in bold.

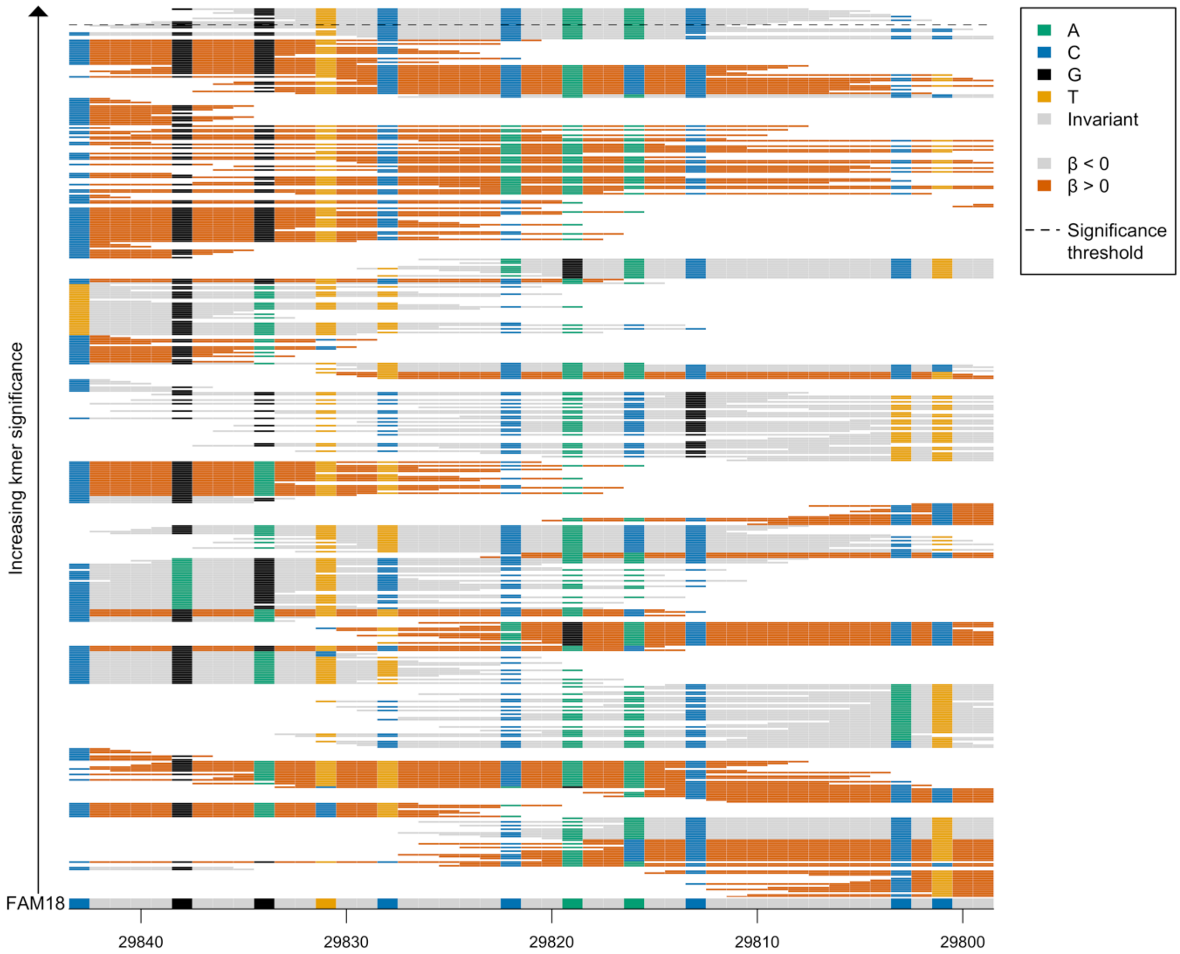


**Supplementary Figure 4** Close up of the significant kmers aligned to the gene *tspB* in the discovery sample collection. The reference genome FAM18 is shown at the bottom of the figure, grey for invariant sites and coloured at variant site positions. The kmers which map to the region shown are then plotted from least significant at the bottom to most significant at the top. The black dashed line indicates the Bonferroni-corrected significance threshold – all kmers above the line are significantly associated with the phenotype. The background colour of the kmers represents the direction of the association, grey when $\beta < 0$ (carriage-associated) and dark orange when $\beta > 0$ (disease-associated). Kmers are coloured by their allele at all variant positions (A = green; C = blue; G = black; T = Orange).

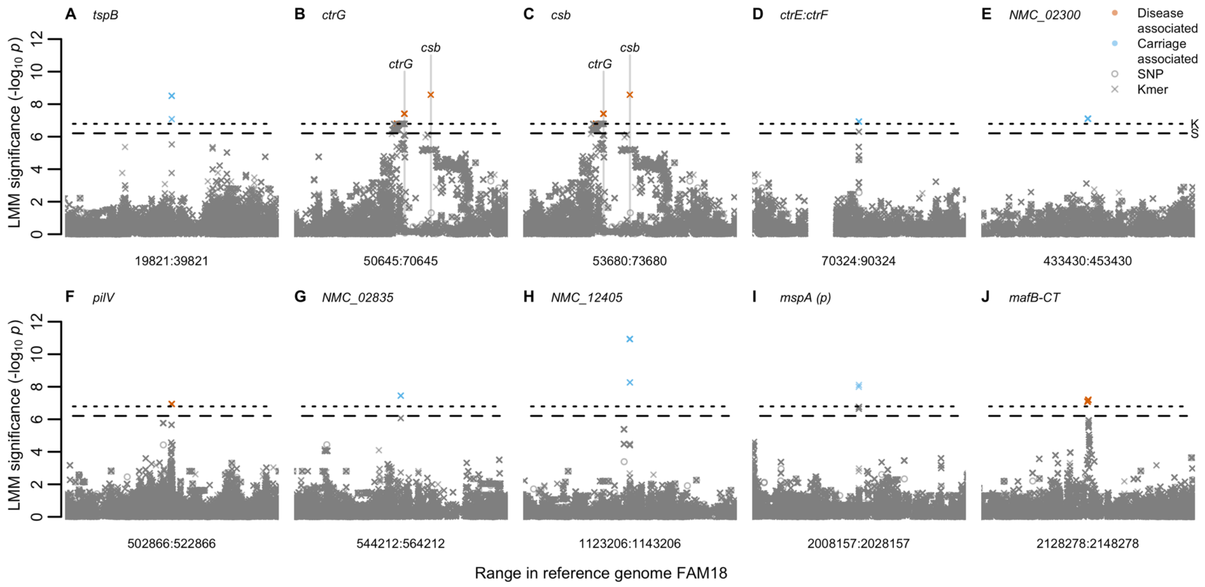


**Supplementary Figure 5** Genes or intergenic regions containing only significant kmers associated with carriage *versus* invasive disease in 261 isolates sampled from the Czech Republic in 1993. Each plot shows the midpoint of the kmer association within the gene/intergenic region +/- 10kb. Open circles represent a SNP aligned to the reference genome FAM18, a cross represents the left-most mapping position of a kmer in the reference genome FAM18 based on mapping and BLAST alignments. Significant kmers are coloured by the LMM estimated direction of effect. Bonferroni-corrected significance thresholds are shown by black dashed (SNPs) and dotted (kmers) lines. Gene names separated by colons indicate intergenic regions. FAM18 reference genome gene name prefixes have been shortened from NMC_RS to NMC_.

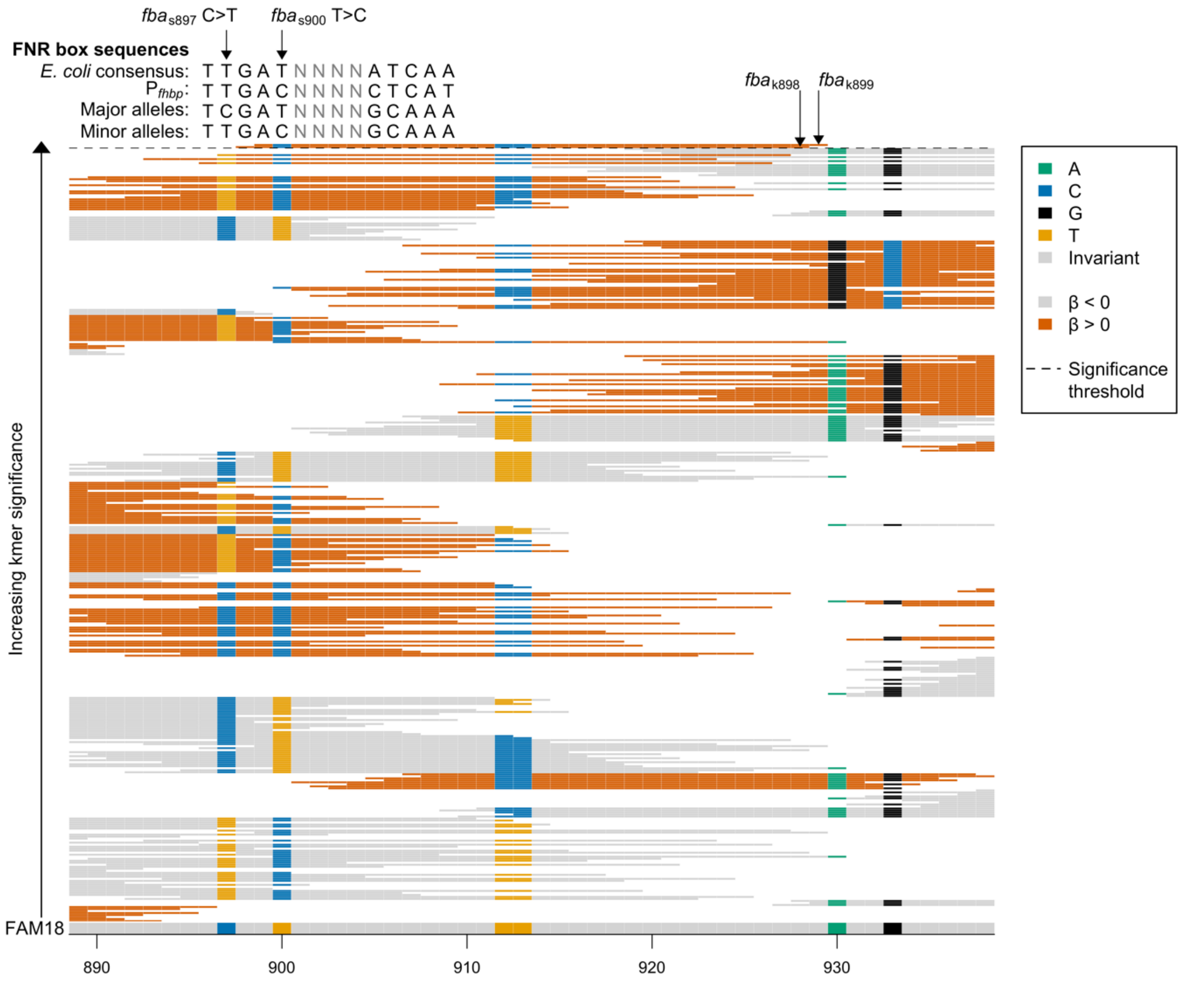


**Supplementary Figure 6** Close up of the significant kmers aligned to the gene *fba* in the discovery sample collection. The reference genome FAM18 is shown at the bottom of the figure, grey for invariant sites and coloured at variant site positions. The kmers which map to the region shown are then plotted from least significant at the bottom to most significant at the top. The black dashed line indicates the Bonferroni-corrected significance threshold – all kmers above the line are significantly associated with the phenotype. The background colour of the kmers represents the direction of the association, grey when $\beta < 0$ (carriage-associated) and dark orange when $\beta > 0$ (disease-associated). Kmers are coloured by their allele at all variant positions (A = green; C = blue; G = black; T = Orange). The *E. coli* consensus for the FNR box DNA binding site and the *fHbp* promoter FNR binding site are shown above the kmers aligned with the major and minor alleles in the discovery sample collection at these positions.

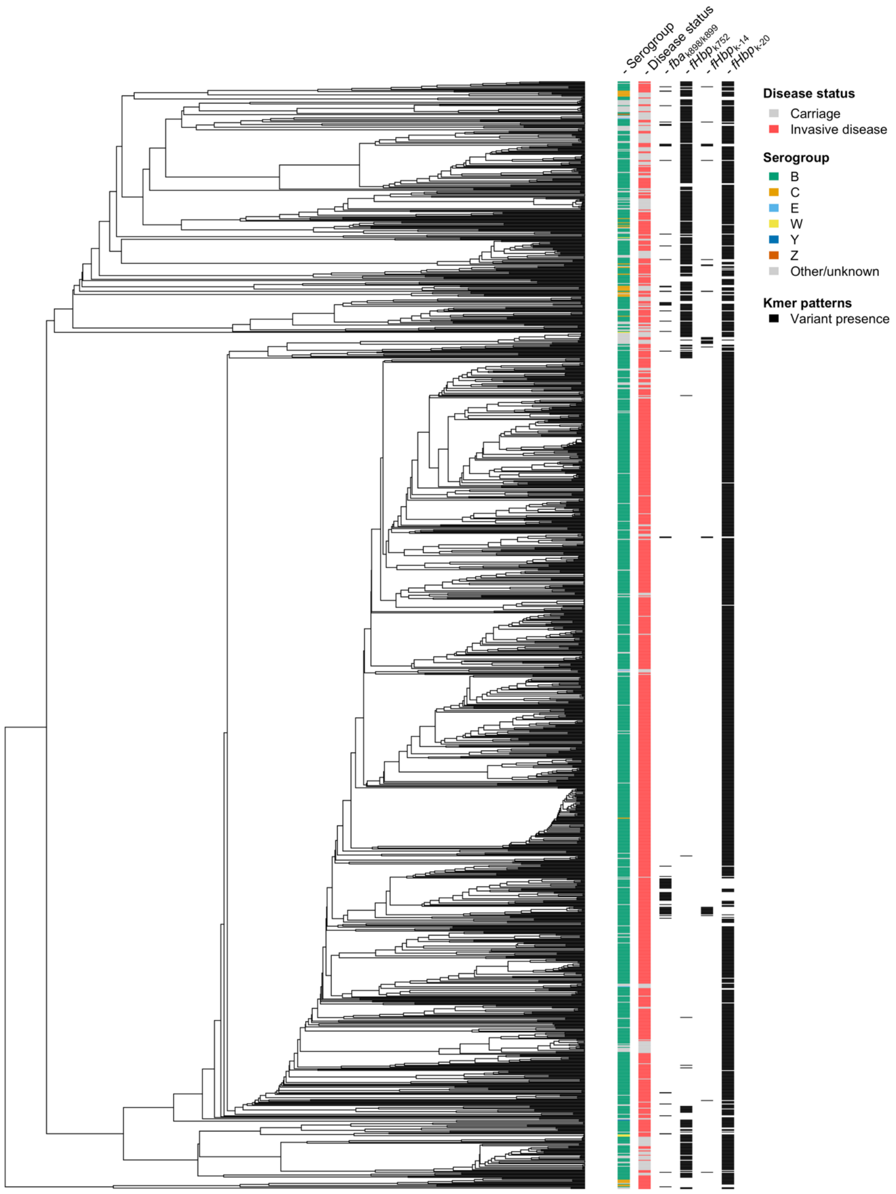


**Supplementary Figure 7** UPGMA tree of 1,295 ST-41/44 complex *N. meningitidis* genomes downloaded from pubMLST. The UPGMA tree, used solely for visualisation, was estimated using a distance matrix calculated from the kmer presence/absence matrix. The most common serogroups are shown on the outer ring. Disease status is shown on the next ring, invasive disease (red, n = 1,046) or carriage (grey, n = 249). Presence of the two significant kmers in *fba* in the discovery sample collection are shown in black in the inner ring.

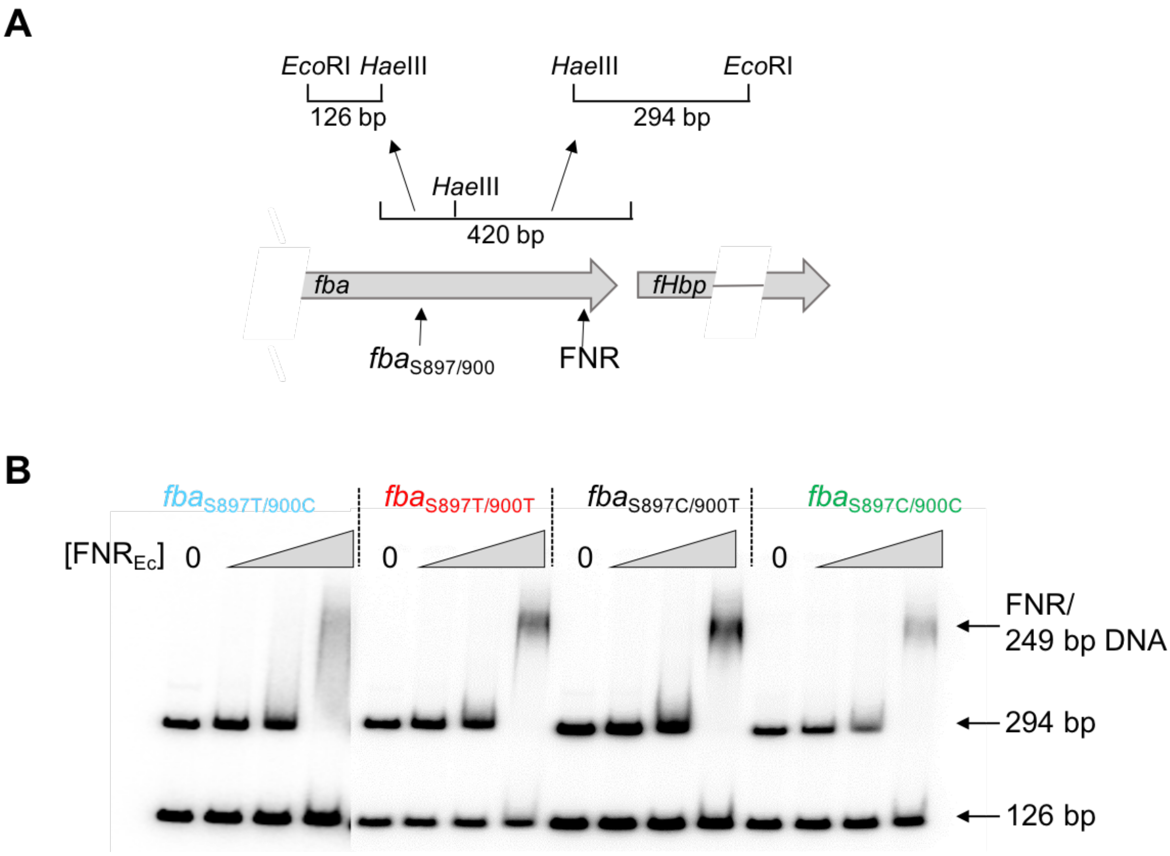


**Supplementary Figure 8 (A)** The 126 and 249 bp fragments used for EMSA. **(B)** Sequences upstream of *fHbp* were amplified and digested with *Hae*III, end labelled with [γ-$^{32}$P]-ATP, then incubated in increasing concentrations of FNR (0, 0.75, 1.5, and 3 µM).

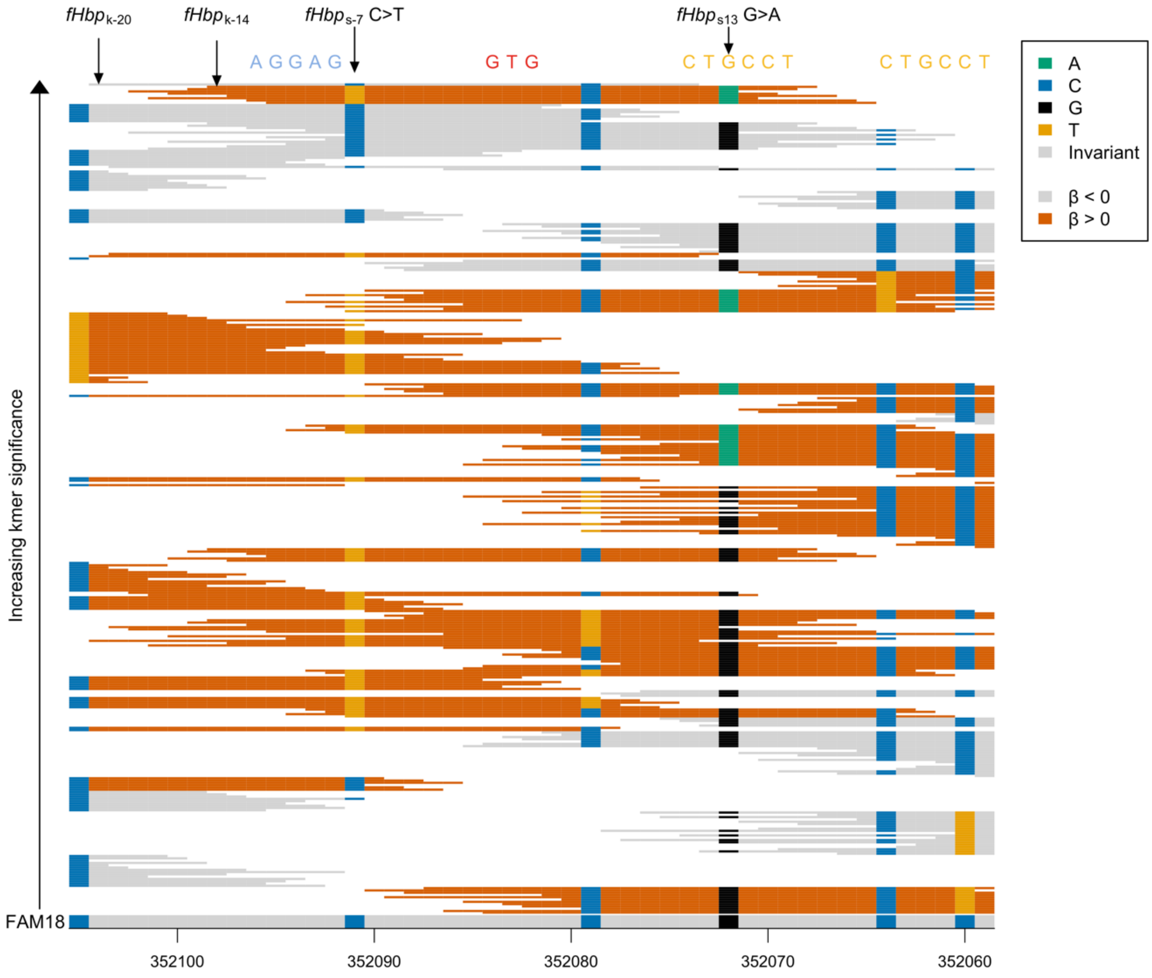


**Supplementary Figure 9** Close up of the ST-41/44 complex replication analysis kmers aligned to the start codon of *fHbp* and the surrounding region. The reference genome FAM18 is shown at the bottom of the figure, grey for invariant sites and coloured at variant site positions. The kmers above 1% minor allele frequency that map or align to the region shown are plotted from least significant at the bottom to most significant at the top. The background colour of the kmers represents the direction of the association, grey when $\beta < 0$ (carriage-associated) and dark orange when $\beta > 0$ (disease-associated). Kmers are coloured by their allele at all variant positions (A = green; C = blue; G = black; T = Orange). The *fHbp* start codon is shown aligned above the kmers in red, the ribosome binding site (RBS) in blue and two putative anti-RBSs (Loh *et al.*, 2016) in orange. The most significant kmers plus the SNPs tested experimentally are labelled.

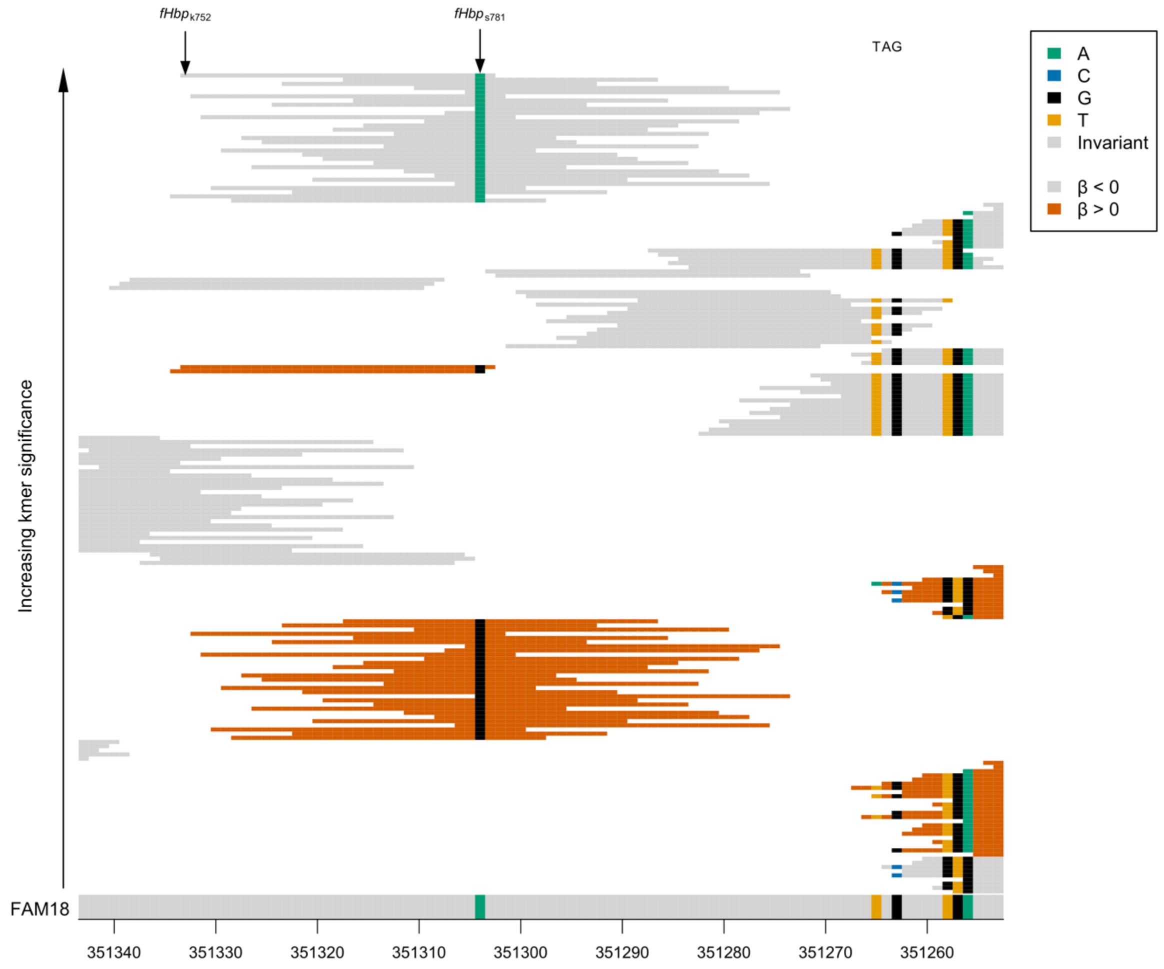


**Supplementary Figure 10** Close up of the ST-41/44 complex replication analysis kmers aligned to the *fHbp* stop codon and upstream region. The reference genome FAM18 is shown at the bottom of the figure, grey for invariant sites and coloured at variant site positions. The kmers which map to the region shown are then plotted from least significant at the bottom to most significant at the top. The background colour of the kmers represents the direction of the association, grey when $\beta < 0$ (carriage-associated) and dark orange when $\beta > 0$ (disease-associated). Kmers are coloured by their allele at all variant positions (A = green; C = blue; G = black; T = Orange). The *fHbp* stop codon is annotated above the aligned kmers. The most significant kmer plus the SNP tested experimentally are labelled.

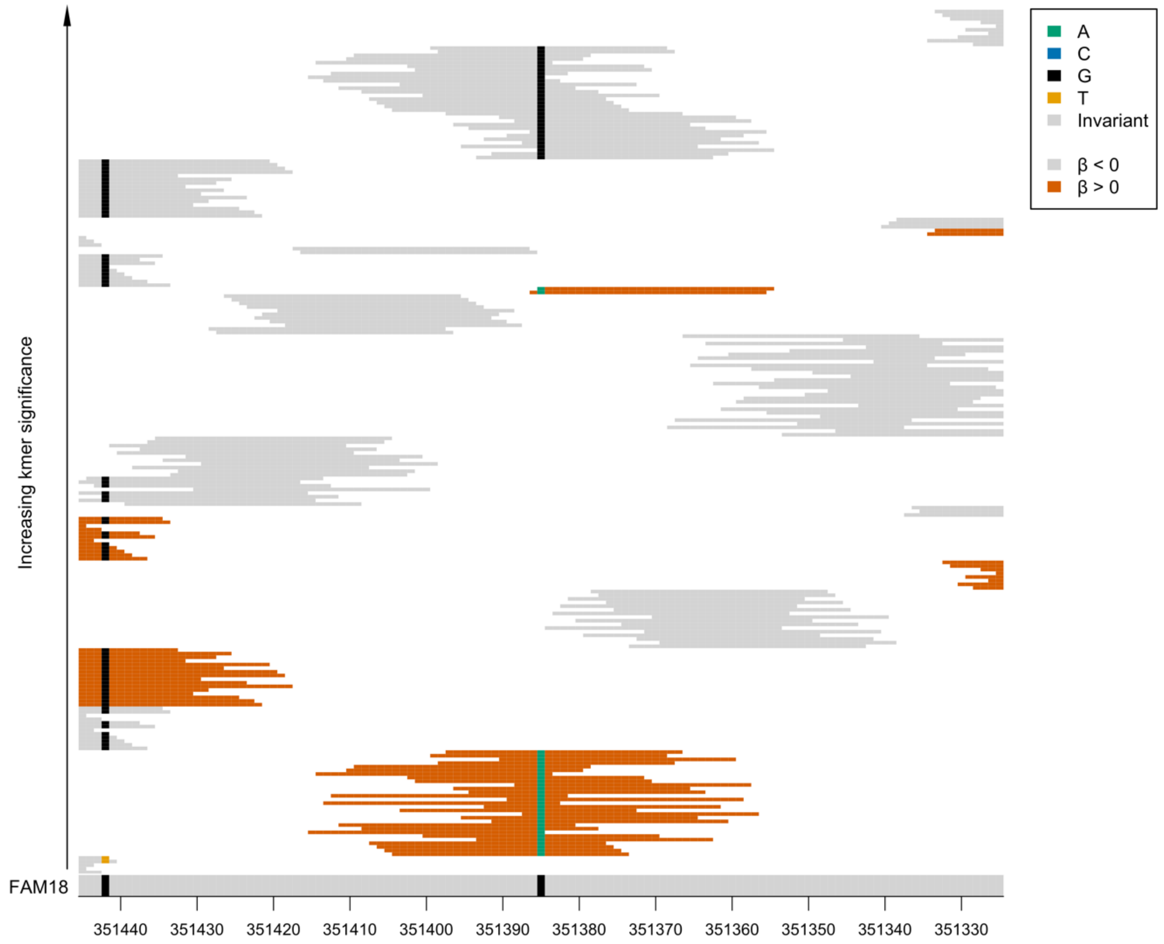


**Supplementary Figure 11** Close up of the ST-41/44 complex replication analysis kmers aligned to the *fHbp* positions 643-760. The reference genome FAM18 is shown at the bottom of the figure, grey for invariant sites and coloured at variant site positions. The kmers which map to the region shown are then plotted from least significant at the bottom to most significant at the top. The background colour of the kmers represents the direction of the association, grey when $\beta < 0$ (carriage-associated) and dark orange when $\beta > 0$ (disease-associated). Kmers are coloured by their allele at all variant positions (A = green; C = blue; G = black; T = Orange). The *fHbp* stop codon is annotated above the aligned kmers in red.

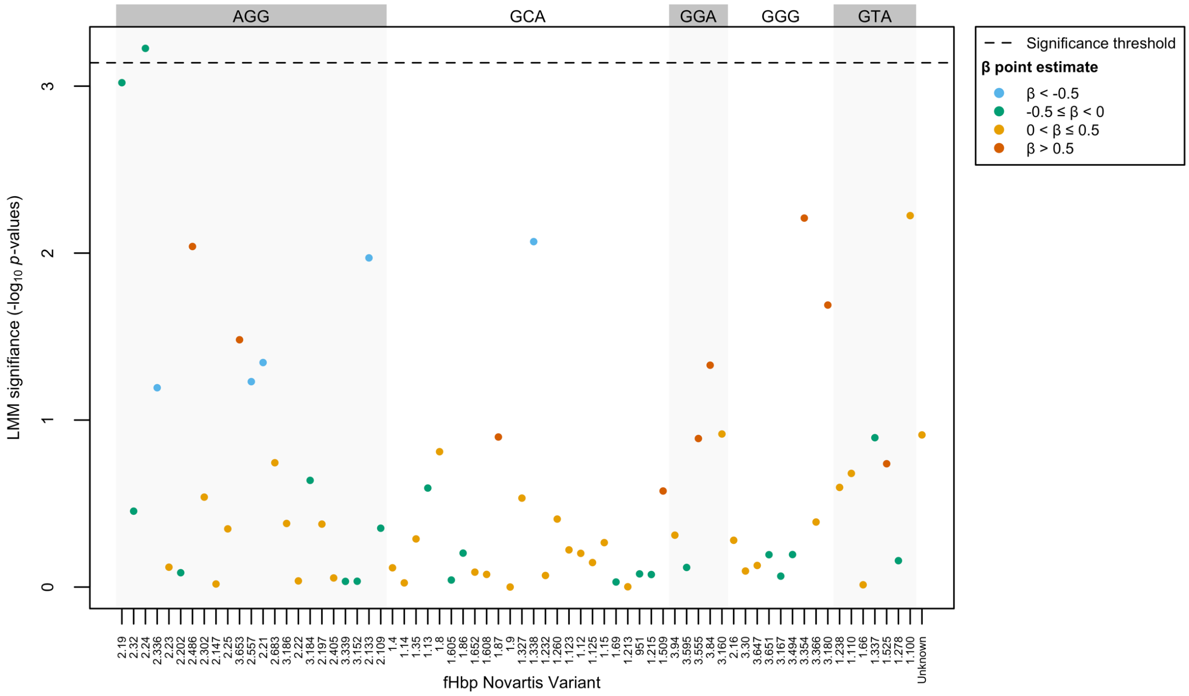


**Supplementary Figure 12** We tested *fHbp* Novartis variants from pubMLST for their association with carriage *vs.* IMD by LMM, using the full kmer kinship matrix to control for population structure. Correcting the significance threshold for the number of variants, variant 2.24 was significantly associated with disease status. The codon each variant contains at codon 261 (relative to the FAM18 reference fHbp) is shown at the top. Colour represents the β point estimate, the direction of the effect of the association by the LMM.

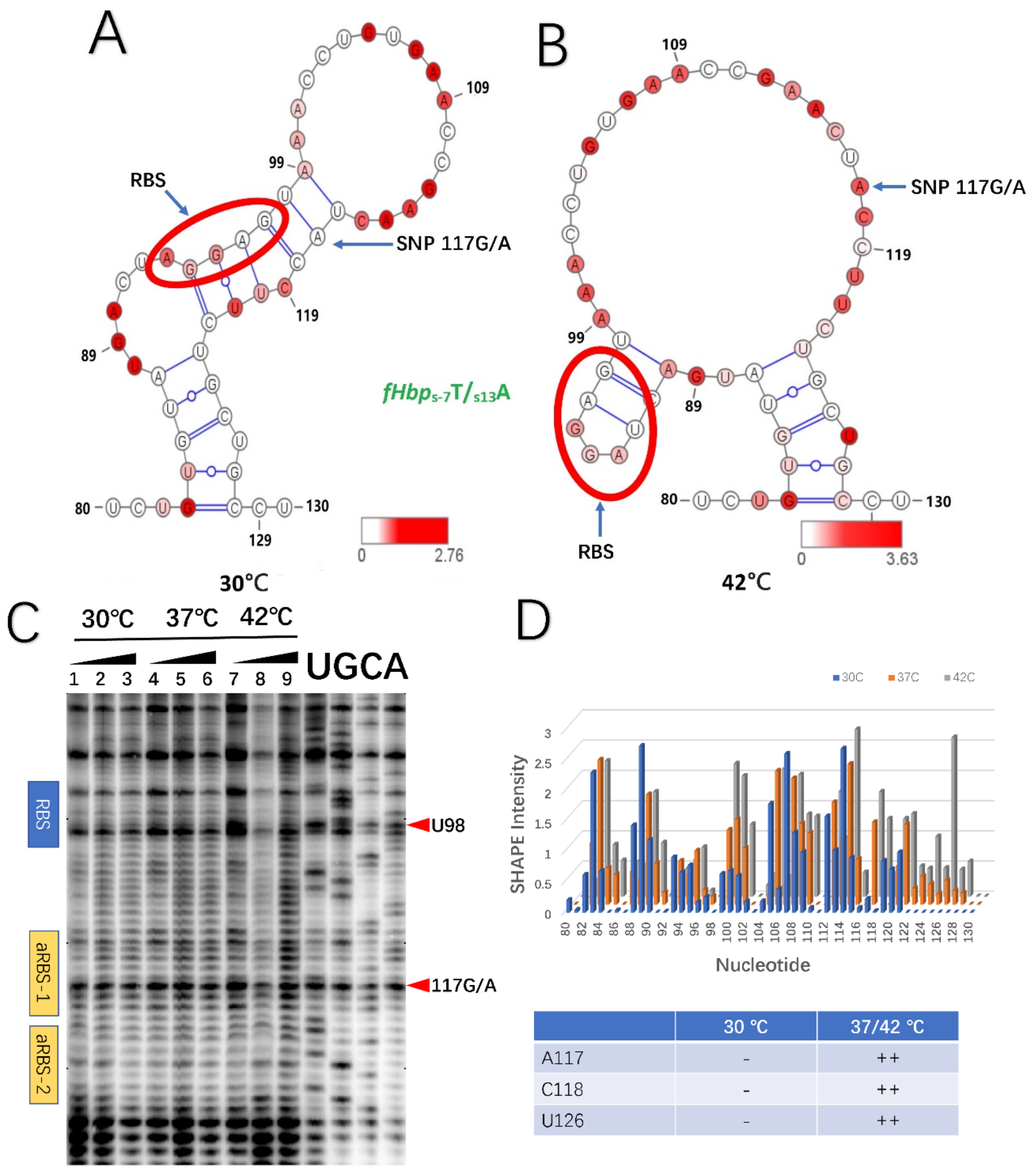


| | 30 °C | 37/42 °C |
|---|---|---|
| A117 | - | ++ |
| C118 | - | ++ |
| U126 | - | ++ |

**Supplementary Figure 13** Secondary structure of *fHbp*s-7T/s13A RNA calculated using RNA structure based on SHAPE reactivity data at **(A)** 30°C and **(B)** 42°C; SHAPE reactivity data are mapped on the RNA structure and colour coded by intensity as shown on the bars; the RBS is circled in red. **(C)** NMIA modifications for reactions conducted at 30°C (Lane1- 3); 37°C (lane 4-6) and 42°C (lane 7-9), with a gradient of 0-13mM NMIA, analysed by denaturing polyacrylamide gel electrophoresis. **(D)** SHAPE reactivity profile at different temperatures; nucleotides with temperature dependent changes in reactivity are listed in the table as strong (++), medium (+) and weak (-).

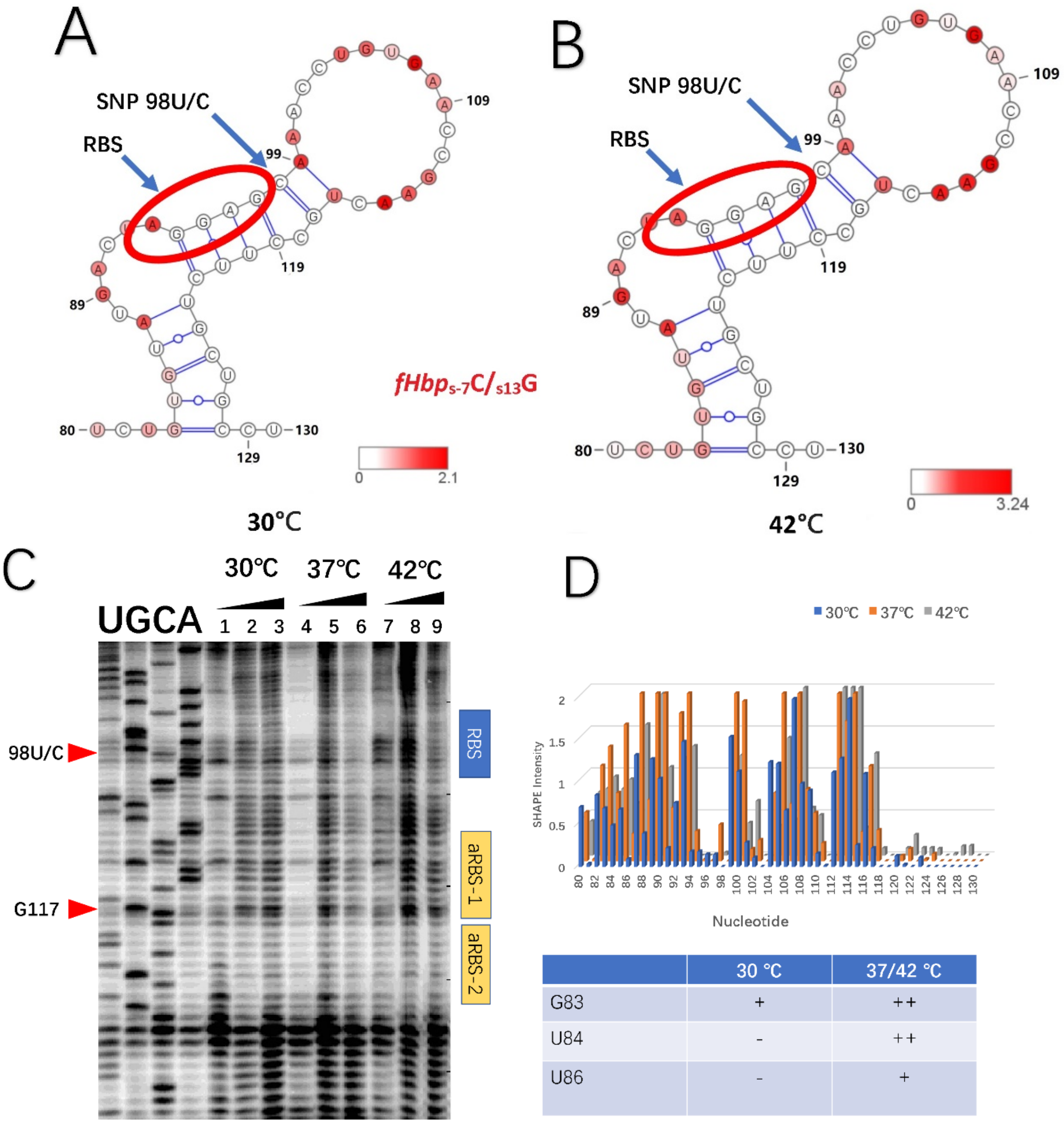


| | 30 °C | 37/42 °C |
|---|---|---|
| G83 | + | ++ |
| U84 | - | ++ |
| U86 | - | + |

**Supplementary Figure 14** Secondary structure of the *fHbp*s-7C/s13G RNA calculated using RNA structure based on SHAPE reactivity data at **(A)** 30°C and **(B)** 42°C; SHAPE reactivity data are mapped on the RNA structure and colour coded by intensity as shown on the bars; the RBS is circled in red. **(C)** NMIA modification for reactions conducted at 30°C (Lane 1-3); 37°C (lane 4-6) and 42°C (lane 7-9), with a gradient of 0-13mM NMIA, analysed by denaturing polyacrylamide gel electrophoresis. **(D)** SHAPE reactivity profile at the different temperatures; nucleotides with temperature dependent changes in reactivity are listed in the table as strong (++), medium (+) and weak (-).

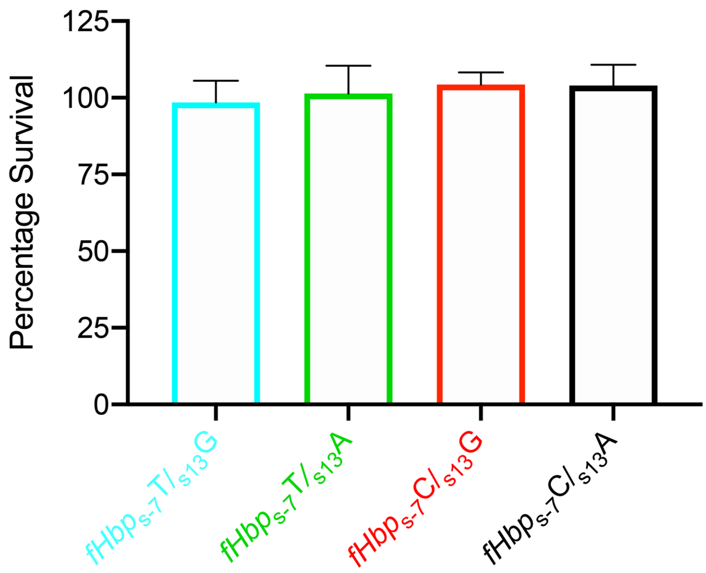


**Supplementary Figure 15** Serum sensitivity assays of *N. meningitidis* strains using heat-inactivated serum. SNPs; *fHbp*$_{s-7}$T/*fHbp*$_{s13}$G (blue), *fHbp*$_{s-7}$T/*fHbp*$_{s13}$A (green), *fHbp*$_{s-7}$C/*fHbp*$_{s13}$G (red) and *fHbp*$_{s-7}$C/*fHbp*$_{s13}$A (black) demonstrated no statistical difference in bacterial survival. Error bars show SD (n=3) with statistical analysis performed in Prism v10 using One-way ANOVA.

**SUPPLEMENTARY TABLES**

**Supplementary Table 1: Summary of significant kmer associations**

The number of significant kmers, most significant -$\log_{10}$ *P* values and $\beta$ point estimates for the most significant kmers for each gene. (p) denotes pseudogenes.

| **Gene(s)** | **No. of significant kmers** | **-$\log_{10}$ *P* values** | **$\beta$ point estimates** |
|---|---|---|---|
| *tspB* | 9 | 8.51 | -0.49 |
| *ctrG* | 45 | 7.41 | 0.43 |
| *csb* | 21 | 8.58 | 0.45 |
| *ctrE-ctrF* intergenic region | 9 | 6.93 | -0.54 |
| *gidA* | 31 | 7.1 | -0.59 |
| *fba* | 2 | 7.16 | 0.59 |
| NMC_RS02300 | 31 | 7.1 | -0.59 |
| *pilV* | 25 | 6.94 | 0.72 |
| frpC operon | 4 | 7.45 | -0.48 |
| *trkH* | 62 | 7.1 | 0.59, -0.59 |
| NMC_RS11425 (p)-*vacB* intergenic region | 48 | 7.92 | 0.58 |
| NMC_RS12405 (p) | 22 | 10.93 | -0.59 |
| NMC_RS06715 | 31 | 7.1 | -0.59 |
| *mutL* | 31 | 7.1 | -0.59 |
| *lptF* | 39 | 7.1 | -0.59, 0.59 |
| *mspA* (p) | 2 | 8.11 | -0.50 |
| *mafB* | 53 | 7.2 | 0.39 |

**Supplementary Table 2: Strains used to test *fba* and *fHbp* SNPs**

| Species/strain | Genotype/Description | Source |
|---|---|---|
| *E. coli* | | |
| Dh5α | F– *endA1 glnV44 thi-1 recA1 relA1 gyrA96 deoR nupG purB20* φ80d*lacZ*ΔM15 Δ(*lacZYA-argF*)U169, hsdR17($r_{K-}m_{K+}$), λ– | Invitrogen |
| *N. meningitidis* | | |
| 0011/93 | Serogroup B, ST-41/44, IMD 1993 | [ref] |
| OX99.32412 | Serogroup C, ST-41/44, Carriage, 1999 | |
| *fHbp*$_{S-7}$T/$_{S13}$G | 0011/93 with *fHbp*$_{S-7}$T/$_{S13}$G, ery$^{R}$ downstream of *fHbp* | This study |
| *fHbp*$_{S-7}$T/$_{S13}$A | 0011/93 with *fHbp*$_{S-7}$T/$_{S13}$A, ery$^{R}$ downstream of *fHbp* | This study |
| *fHbp*$_{S-7}$C/$_{S13}$G | 0011/93 with *fHbp*$_{S-7}$C/$_{S13}$G, ery$^{R}$ downstream of *fHbp* | This study |
| *fHbp*$_{S-7}$C/$_{S13}$A | 0011/93 with *fHbp*$_{S-7}$C/$_{S13}$A, ery$^{R}$ downstream of *fHbp* | This study |

**Supplementary Table 3: Plasmids used in this study**

| Plasmid | *E. coli* | Description | Source |
|---|---|---|---|
| pET28a | DH5α | - | Novagen |
| pET21b | DH5α | - | Novagen |
| pET24-HIS-TEV | DH5α | TEV protease cleavage site tagged with HIS | This study |
| pET28a-His-MBP-TEV | DH5α | For expression of fHbps fused to MBP fusion | This study |
| pET21b_V2.24_A | B834 | vector for IPTG inducible expression of fHbp V2.24$^{261R}$ | This study |
| pET21b_V2.24_G | B834 | vector for IPTG inducible expression of fHbp V2.24$^{261G}$ | This study |
| pET21b_V1.1$^{I311A}$ | B834 | vector for IPTG inducible expression of fHbp V1.1$^{I311A}$ | Johnson *et al.* (2013) |
| pET28a_V2.24_A | B834 | vector for IPTG inducible expression of fHbp V2.24$^{261R}$ | This study |
| pET28a_V2.24_G | B834 | vector for IPTG inducible expression of fHbp V2.24$^{261G}$ | This study |
| pUC19 | DH5α | Cloning vector | This study |
| pUC19*fHbp*$_{S-7}$T/$_{S13}$G | DH5α | Generating construct | This study |
| pUC19*fHbp*$_{S-7}$T/$_{S13}$A | DH5α | Generating construct | This study |
| pUC19 *fHbp*$_{S-7}$C/$_{S13}$G | DH5α | Generating construct | This study |
| pUC19 *fHbp*$_{S-7}$C/$_{S13}$A | DH5α | Generating construct | This study |
| pUC19*fba*$_{S897}$T/$_{s900}$C | DH5α | Generating construct | This study |
| pUC19*fba*$_{S897}$C/$_{s900}$C | DH5α | Generating construct | This study |
| pUC19*fba*$_{S897}$T/$_{s900}$T | DH5α | Generating construct | This study |
| pUC19*fba*$_{S897}$C/$_{s900}$T | DH5α | Generating construct | This study |

**Supplementary Table 4: Primers used in this study**

| Name | Sequence |
|---|---|
| pET28a-HIS-MBP-TEV-F | ACTTTAAGAAGGAGATATACCATGGGCCATCACCATCACCATC |
| pET28a-HIS-MBP-TEV-R | CGAGTGCGGCCGCAAGCTTGTCGACCCTGAAAATACAGATTTTCGCTACCCGGAGT CTGCGC |
| pET28a-GST-F | CTGGTGCCGCGCGGCAGCCATATGTCCCCTATACTAGGTTATTG |
| pET28a-GST-R | AGCTTTCTCCTTTTGGAGGATGGTCGC |
| pET28a-TEV-F | TCCTCCAAAAGGAGAAAGCTTGTTTAAGG |
| pET28a-TEV-R | GTCGACGGAGCTCGAATTCGGATCCTTAAGAACCAGGTTCTTC |
| pET21b_2.24_SNP_F | GCcatatgatggccgccgacatCGGCGCGGGGCTT |
| pET21b_2.24_SNP_A_R | GCctcgagctgtttgccggcGATGCCGATTTCGTGAACCTTTTCCCTTATCTTCACGGTTGC |
| pET21b_2.24_SNP_G_R | GCctcgagctgtttgccggcGATGCCGATTTCGTGAACCTTTTCCCCTATCTTCACGGTTGC |
| V2fHbp_pET28a_MBP_Tev_F | aaatctgtattttcagggtGTTGCCGCCGACATCGGCG |
| V2fHbp_ pET28a_MBP_Tev_R | agtggtggtggtggtggtgctcgagTTACTGTTTGCCGGCGATGCC |
| ML428 | GATCCTCTAGAGTCGACCTGCAGGCATGCAAAAGTCATCAACGAATATGGC |
| ML429 | AGAAACGAATCTGTATTTTAATTTGTCCGAGGGCGGTATGGCGCAAAAATG |
| ML430 | TCGGACAAATTAAAATACAGATTCG |
| ML435 | GCCGTGCCGTCGTGTCCTGGTACACGAAAAACAAGTTAAG |
| ML433 | CAGGAAACAGCTATGACCATGATTACGCCAGGGCGATTTTGTTGCGGACG |
| ML434 | CTTGTTTTTCGTGTACCAGGACACGACGGCACGGC |
| ML436 | GTGAACCGAACTACCTTCTGCTGCCTTTTCCTGAC |
| ML437 | GAAAAGGCAGCAGAAGGTAGTTCGGTTCACAGGTTTACTC |
| ML438 | ATGACTAGGAGCAAACCTGTGAACCGAACTGC |
| ML439 | AGTTCGGTTCACAGGTTTGCTCCTAGTCATACACAGAATAG |

| | |
|---|---|
| ML440 | TAGGAGCAAACCTGTGAACCGAACTACCTTCTGCTGCCTTTTCCTG |
| ML441 | AAGGTAGTTCGGTTCACAGGTTTGCTCCTAGTCATACACAGAATAG |
| ML405 | TCCCGGCAACAATTAATAGAC |
| ML406 | CCAGTCTATTAATTGTTGCCGG |
| ERS001 | cgacgttgtaaaacgacggccagtgaattcCCGTGGTTAATTTCTCCCAC |
| ERS002 | ggtatttgcgcggaTCAAAGTCGGACGGGTTTTC |
| ERS003 | cgtccgactttgatCCGCGCAAATACCTGAGC |
| ERS004 | tcgatgatggttgGGCGCAAAAATGTTACTGTTTG |
| ERS005 | acatttttgcgccCAACCATCATCGATGAATTGTG |
| ERS006 | cctagggcggtatCGATGCATGCCAACAGATAAAAC |
| ERS007 | ttggcatgcatcgATACCGCCCTAGGACACGAC |
| ERS008 | aagcttgcatgcctgcaggtcgactctagaGGGCAATGTCTGCCGCCC |
| ERS009 | ATTTGCGCGGGTCGAAGTCGGACGGGTTTTCG |
| ERS010 | CGTCCGACTTCGACCCGCGCAAATACCTGAGC |
| ERS011 | ATTTGCGCGGATCGAAGTCGGACGGGTTTTCG |
| ERS012 | TCGGTTCACAGGTTTACTCCTAGT |
| ER013 | CAAAGTCAACATCGACACCGACC |
| GV1 | TTCCGCCGCCTCCGCTGCTGCA |
| GV2 | GCGGTCAGAATCAGGGCGGTGG |
| GV3 | GGTTTACTCCTAGTCATACACAGA |

**Supplementary Table 5: Sequences used for SHAPE analysis**

| Name | Sequence |
|---|---|
| *fHbp*$_{s-7}$T/<br>*fHbp*$_{s13}$G | UUUUUGCUUC UUUGACCUGC CUCAUUGAUG CAAUAUGCAA AAAAAGAUAC<br>CGCAACCAAA ACGUUUAUAU AUUAUCUAUU CUGUGUAUGA CUAGGAGUAA<br>ACCUGUGAAC CGAACUGCCU UCUGCUGCCU UUUCCUGACC ACCGCCCUGA<br>UUCUGACCGC CUGCAGCAGC GGAGGCGGCG GAA |
| *fHbp*$_{s-7}$T/<br>*fHbp*$_{s13}$A | UUUUUGCUUC UUUGACCUGC CUCAUUGAUG CAAUAUGCAA AAAAAGAUAC<br>CGCAACCAAA ACGUUUAUAU AUUAUCUAUU CUGUGUAUGA CUAGGAGUAA<br>ACCUGUGAAC CGAACUACCU UCUGCUGCCU UUUCCUGACC ACCGCCCUGA<br>UUCUGACCGC CUGCAGCAGC GGAGGCGGCG GAA |
| *fHbp*$_{s-7}$C/<br>*fHbp*$_{s13}$G | UUUUUGCUUC UUUGACCUGC CUCAUUGAUG CAAUAUGCAA AAAAAGAUAC<br>CGCAACCAAA ACGUUUAUAU AUUAUCUAUU CUGUGUAUGA CUAGGAGCAA<br>ACCUGUGAAC CGAACUGCCU UCUGCUGCCU UUUCCUGACC ACCGCCCUGA<br>UUCUGACCGC CUGCAGCAGC GGAGGCGGCG GAA |
| *fHbp*$_{s-7}$C/<br>*fHbp*$_{s13}$A | UUUUUGCUUC UUUGACCUGC CUCAUUGAUG CAAUAUGCAA AAAAAGAUAC<br>CGCAACCAAA ACGUUUAUAU AUUAUCUAUU CUGUGUAUGA CUAGGAGCAA<br>ACCUGUGAAC CGAACUACCU UCUGCUGCCU UUUCCUGACC ACCGCCCUGA<br>UUCUGACCGC CUGCAGCAGC GGAGGCGGCG GAA |

### SUPPLEMENTARY INFORMATION

The following contains further detail on the nature of the kmer GWAS associations discussed in the main text and the type of variation that we believe the kmers were capturing.

#### Supplementary text 1

**Phase variable region in *csb*.** There are two phase variable regions in the serogroup B *csb* coding sequence, a poly(C) homopolymeric tract and a poly(A) homopolymeric tract [1, 2]. The 21 significant disease-associated kmers in *csb* covered a poly(A) phase variable homopolymeric tract within the coding sequence, previously described in serogroup B isolate MC58 (NC_003112.2) *csb* gene and all contained poly(A) tract lengths of 9. The remaining isolates containing the serogroup B *csb* allele contained poly(A) tract lengths of 8, 10 or 11. We bioinformatically predicted *csb* state, whether phase variable "on" or "off", determined by whether the assembled gene contained frameshifts or premature stop codons relative to the serogroup B isolate MC58. Of the 68 isolates containing the significant kmers, the gene was split over two contigs for one isolate, for the remainder 63/68 were predicted to be "on" as they did not contain any frameshifts or premature stop codons. 4/68 contained an insertion in the poly(C) phase variable region and therefore a frameshift and premature stop codon so were predicted to be "off". All serogroup B *csb* allele positive isolates for which the significant kmers were absent contained premature stop codons. Presence of the significant *csb* kmers therefore appeared to be representing the serogroup B *csb* "on" state within the poly(A) phase variable region.

**Putative promoter in the intergenic region between *ctrE* and *ctrF*.** Nine significant carriage-associated kmers mapped to the intergenic region between the two genes and contained the major alleles of three SNPs in the region (**Supplementary Figure 3**). The *ctrF* transcriptional start site has been mapped and the -10 box upstream of the gene predicted according to consensus sequences, although no -35 box was detected [3]. We hypothesise that the kmers cover the promoter region for *ctrF* and that divergence from the kmer sequences effects the expression of *ctrF*. The sequence covered by the significant kmers reveal a match to four of the six nucleotides of the *E. coli* consensus sequence for the $\sigma^{70}$ -10 Pribnow box and a match to three out of six nucleotides for the -35 sequence, separated by 17 bp, the optimal promoter spacing. The *E. coli* consensus is 5'-**TTG**ACA-17 bp-**TA**TAA**T**-3' [4] and the sequence captured by the kmers is 5'-TTGCAT-17 bp-TATGCT-3', with the most conserved *E. coli* bases shown in bold. The intergenic region therefore contains the most highly conserved residues in *E. coli*, separated by 17 bp the optimal spacing for an actively transcribed promoter.

The discovery sample collection contained three SNPs within the 17 bp separating the putative -10 and -35 promoter regions, at positions -14, -19 and -28 (**Supplementary Figure 3**). The carriage-associated major alleles for these SNPs were G, C and C, and the second most common IMD-associated alleles were all Ts. It has previously been shown that mutating the $P_{lac}$ promoter spacer sequence from GC-rich to AT-rich made it hyperactive [5]. Also, the SNP at position -19 is 1 bp away from position -18, the previously detected most important T residue in the *E. coli* spacer region for stimulating transcription [6]. Two of the SNP T alleles increase the length of poly(T) tracts and it has been shown that the extent of promoter DNA bending in an *E. coli* promoter was proportional to the number of $T_5$ and $T_6$ tracts in the promoter [7]. It has also been found that an *E. coli* promoter spacer sequence influenced promoter activity and the curvature of the DNA [8]. We therefore hypothesise that the SNPs could affect DNA curvature in the region and therefore expression of *ctrF*.

Many bacterial species including *N. meningitidis* contain an extended -10 promoter, which is a '5-TRTG-3' sequence (where R is A or G) positioned one nucleotide upstream of -10 box [3, 9, 10]. The extended -10 region has been shown to strengthen the promoter activity by enhancing the interaction between the promoter and RNA polymerase-sigma factor transcription initiation complex [11]. Mutagenesis analysis of *E. coli* promoters has revealed that substitutions in extended -10 region differentially affect promoter activity [12]. Interestingly, *ctrF* promoter contains a putative extended -10 sequence (**Supplementary Figure 3**), which harbours the SNP at position -14. Therefore, in addition to two SNPs in the spacer region of *ctrF* promoter, -14 SNP may further impact promoter activity and expression of *ctrF*.

**Meningococcal Disease Associated Island gene *tspB*.** Nine significant kmers were identified in *tspB*, one of the genes of the MDA phage. The effect of *tspB* deletions in the isolate H44/76, which has three complete copies of *tspB*, has been tested and found that one copy (nmbh4476_0681) was more important than the other two for resistance to normal human serum (NHS); the mutant solely carrying nmbh4476_0681 was as resistant to NHS as the parent strain [13]. The significant kmers match perfectly to this copy of *tspB* and not to the other two complete copies in H44/76. The region of *tspB* that the kmers map to is the amino-terminal domain of the highly conserved region, which is the domain of TspB which binds to IgG. The significant kmers altogether cover nine SNPs, of these two differentiate the H44/76 *tspB* copy important for resistance to NHS from the other two copies which are both synonymous SNPs. Therefore the kmers are possibly capturing the H44/76 nmbh4476_0681 allele of *tspB*.

**Supplementary text 2**

The *fHbp* promoter is regulated by FNR (fumarate and nitrate reduction regulator) in response to anaerobiosis [14]. The FNR binding site in the model organism *E. coli* is a highly conserved 5 bp inverted repeat separated by 4 pairs of non-specific bases (TTGAT-$N_4$-ATCAA) [15]. A putative FNR box was discovered in the intergenic region upstream of *fHbp* in *N. meningitidis*, based on experiments which discovered the start point of the *fHbp* mRNA and found that FNR bound a motif overlapping the -35 box of the *fHbp* promoter [14]. The motif differed from the *E. coli* consensus by three nucleotides (TTGAC-$N_4$-CTCAT, **Supplementary Figure 6**).

The disease-associated alleles of the most-significant discovery SNPs *fba*900 and *fba*897 formed the motif (TTGAC-$N_4$-GCAAA), whose first pentamer matched that of the FNR-binding site in the *fHbp* promoter ( [14], **Supplementary Figure 6**). This may therefore constitute a second FNR box antagonistic to the FNR box in the promoter. Many FNR-regulated genes contain two FNR binding sites, where the downstream site activates expression and the upstream site downregulates expression. When FNR is activated due to oxygen depletion it occupies the downstream site activating expression. As FNR becomes fully activated a lower affinity upstream site is filled, downregulating expression and providing a mechanism for microaerobic regulation of gene expression [16, 17, 18]. The spacing between and relative affinities of the two FNR sites are crucial for FNR-mediated repression, with the inward-facing subunits being the most important [18]. We therefore hypothesised that the SNPs in *fba* create a second, antagonistic, FNR box, modifying the expression of *fHbp*.

The disease-associated alleles of *fba*900 *and fba*897 were ubiquitous among the ST-11 complex isolates in the Czech study, whereas they coexisted at intermediate frequency with the non-disease associated alleles in the ST-41/44 complex isolates. We therefore created isogenic mutants of the ST-41/44 complex ST-740 strain 0011/93 (pubMLST ID 1955) [19] comprising all four combinations of disease-associated and carriage-associated alleles of *fba*900 and *fba*897. However, we found no differences in expression of *fHbp* between the four mutants, and electrophoresis mobility shift assays did not demonstrate binding of a constitutively active version of FNR to the sequences within *fba*, so we were unable to support the hypothesis that the disease-associated alleles of *fba*900 and *fba*897 form a second FNR box.

### SUPPLEMENTARY REFERENCES


[1] S. Hammerschmidt, A. Müller, H. Sillmann, M. Mühlenhoff, R. Borrow, A. Fox, J. Van Putten, W. D. Zollinger, R. Gerardy-Schahn and M. Frosch, "Capsule phase variation in Neisseria meningitidis serogroup B by slipped-strand mispairing in the polysialyltransferase gene (siaD): Correlation with bacterial invasion and the outbreak of meningococcal disease," *Molecular Microbiology,* vol. 20, no. 6, pp. 1211-1220, 1996.

[2] M. V. R. Weber, H. Claus, M. C. J. Maiden, M. Frosch and U. Vogel, "Genetic mechanisms for loss of encapsulation in polysialyltransferase-gene-positive meningococci isolated from healthy carriers," *International Journal of Medical Microbiology,* vol. 296, no. 7, pp. 475-484, 2006.

[3] N. Heidrich, S. Bauriedl, L. Barquist, L. Li, C. Schoen and J. Vogel, "The primary transcriptome of Neisseria meningitidis and its interaction with the RNA chaperone Hfq," *Nucleic Acids Research,* vol. 45, no. 10, p. 6147–6167, 2017.

[4] C. B. Harley and R. P. Reynolds, "Analysis of E. coli promoter sequences.," *Nucleic Acids Research,* vol. 15, no. 5, pp. 2343-2361, 1987.

[5] M. Liu, M. Tolstorukov, V. Zhurkin, S. Garges and S. Adhya, "A mutant spacer sequence between -35 and -10 elements makes the Plac promoter hyperactive and cAMP receptor protein-independent," *Proceedings of the National Academy of Sciences,* vol. 101, no. 18, pp. 6911-6916, 2004.

[6] S. S. Singh, A. Typas, R. Hengge and D. C. Grainger1, "Escherichia coli σ70 senses sequence and conformation of the promoter spacer region," *Nucleic Acids Research,* vol. 39, no. 12, pp. 5109-5118, 2011.

[7] T. Lozinski, K. Adrych-Rozek, W. T. Markiewicz and K. L. Wierzchowski, "Effect of DNA bending in various regions of a consensuslike Escherichia coli promoter on its strength in vivo and structure of the open complex in vitro," *Nucleic Acids Research,* vol. 19, no. 11, pp. 2947-2953, 1991.

[8] I. G. Hook-Barnard and D. M. Hinton, "The promoter spacer influences transcription initiation via σ70 region 1.1 of Escherichia coli RNA polymerase," *Proceedings of the National Academy of Sciences,* vol. 106, no. 3, pp. 737-742, 2009.

[9] C. P. Moran Jr., N. Lang, S. F. J. LeGrice, G. Lee, M. Stephens, A. L. Sonenshein, P. Janice and R. Losick, "Nucleotide sequences that signal the initiation of transcription and translation in Bacillus subtilis," *Molecular and General Genetics MGG,* vol. 186, no. 3, pp. 339-346, 1982.

[10] M. I. Voskuil, K. Voepel and G. H. Chambliss, "The -16 region, a vital sequence for the utilization of a promoter in Bacillus subtilis and Escherichia coli," *Molecular Microbiology,* vol. 17, no. 2, pp. 271-279, 1995.

[11] M. I. Voskuil and G. H. Chambliss, "The TRTGn motif stabilizes the transcription initiation open complex," *Journal of Molecular Biology,* vol. 322, no. 3, pp. 521-532, 2002.

[12] J. E. Mitchell, D. Zheng, S. J. Busby and S. D. Minchin, "Identification and analysis of 'extended -10' promoters in Escherichia coli," *Nucleic Acids Research,* vol. 31, no. 16, pp. 4689-4695, 2003.

[13] M. G. Müller, N. E. Moe, P. Q. Richards and G. R. Moe, "Resistance of Neisseria meningitidis to Human Serum Depends on T and B Cell Stimulating Protein B," *Infection and Immunity,* vol. 83, no. 4, pp. 1257-1264, 2015.

[14] F. Oriente, V. Scarlato and I. Delany, "Expression of factor H binding protein of meningococcus responds to oxygen limitation through a dedicated FNR-regulated promoter," *Journal of Bacteriology,* vol. 192, no. 3, pp. 691-701, 2010.

[15] K. Eiglmeier, N. Honoré, S. Iuchi, E. C. C. Lin and S. T. Cole, "Molecular genetic analysis of FNR-dependent promoters," *Molecular Microbiology,* vol. 3, no. 7, pp. 869-878, 1989.

[16] J. Guest, J. Green, A. Irvine and S. Spiro, "The FNR modulon and FNR-regulated gene expression," in *Regulation of gene expression in Escherichia coli*, Springer US, 1996, pp. 317-342.

[17] S. M. Williams, H. J. Wing and S. J. W. Busby, "Repression of transcription initiation by Escherichia coli FNR protein: Repression by FNR can be simple," *FEMS Microbiology Letters,* vol. 163, no. 2, pp. 203-208, 1998.

[18] F. A. Marshall, S. L. Messenger, N. R. Wyborn, J. R. Guest, H. Wing, S. J. W. Busby and J. Green, "A novel promoter architecture for microaerobic activation by the anaerobic transcription factor FNR," *Molecular Microbiology,* vol. 39, no. 3, pp. 747-753, 2001.

[19] E. Bille, J.-R. Zahar, A. Perrin, S. Morelle, P. Kriz, K. Jolley, M. C. J. Maiden, C. Dervin, X. Nassif and C. R. Tinsley, "A chromosomally integrated bacteriophage in invasive meningococci.," *J Exp Med,* vol. 201, no. 12, pp. 1905-1913, 2005.
